# Constructing and analyzing a synthetic life course cohort based on pooling two data sources: A case study of early adulthood depression symptomatology and late-life cognition

**DOI:** 10.64898/2026.02.25.26347113

**Authors:** Scott C Zimmerman, Peter Buto, Katrina Kezios, Adina Zeki Al Hazzouri, M Maria Glymour

## Abstract

**Background:** Synthetic cohorts created by combining two cohorts can be useful when no single data set includes both the exposure and outcome data of interest. We estimate the effects of depression in early adulthood on later-life memory outcome using two nationally representative cohorts separately and in a synthetic sample.

**Methods:** We used the National Longitudinal Study of Youth 1979 (NLSY; N=5,747) and the Health and Retirement Study (HRS; N=6,846) and a synthetic cohort combining exposure data from N=5,680 NLSY participants (born 1957-1965) aged 55-63 in 2020 who completed midlife cognitive assessment between 2006-2020 with outcome data from N=9,726 HRS participants born 1957-1964 who completed cognitive assessments when 47-63 years old and every 2-years thereafter. A 6-item version of the Centers for Epidemiologic Studies-Depression (CES-D) score (range 0-6) was measured from late adolescence through midlife in NLSY and in midlife in HRS. Memory was measured as the sum of immediate and delayed word recall scores up to twice in NLSY at age 48+ and up to 10 times in HRS at age 50+. We generated a synthetic life course cohort, matching HRS participants to NLSY participants based on 10 variables measured in midlife in both cohorts and posited to either confound or mediate the association between early life depressive symptoms and late-life memory. Matching variables included midlife depression and memory. We used confounder-adjusted linear mixed models to estimate the association between earliest reported depressive symptoms in NLSY and HRS with memory in the respective data sets and evaluated associations of early life depression symptoms with the repeated later life memory measures in the synthetic cohort.

**Results:** In NLSY, a one point higher CES-D at age 23-31 was associated with a lower (β_NLSY_level_=-0.023 (95%CI:-0.032, −0.015) point difference) average summed recall during follow up (“cognitive level”), but no difference in words recalled per CES-D point per decade of follow-up (“cognitive slope”; β_NLSY_slope_=0.004 (95%CI:-0.034, 0.042) points per decade). In HRS, a one point higher CES-D in midlife (average age 53) was associated with a lower (β_HRS_level_=-0.163 (95%CI:-0.199,-0.128) point difference) average cognitive level but no difference in cognitive slope (β_HRS_slope_=-0.021 (95%CI:-0.062,0.020) points per decade). In the synthetic cohort, a one-point increase in CES-D at age 23-27 was associated with a lower cognitive level (β_Synth_level_=-0.018 (95%CI:-0.030,-0.006) point difference), but no difference in cognitive slope (β_Synth_slope_=0.001 (95%CI:-0.011,0.012) points per decade) at age 50+.

**Conclusions:** Depressive symptoms ages 23-31 predicted mid-to late-life memory function but had no clear association with memory decline. Combining data across cohorts spanning separate, but overlapping, parts of the life course is a promising approach to overcome data limitations in life course research, but it requires careful implementation to ensure that assumptions are met and estimates are appropriately interpreted.

## Introduction

Depression, which affects 8-20% of adults (Goodwin 2022, Jiang 2025, Köse 2025), is associated with approximately a doubling in the risk of dementia (Stafford 2022), and a threefold increase in risk for those diagnosed before age 44 (Elser 2023). Treating midlife depression may reduce lifetime dementia prevalence by 3.0-8.3% (Livingston 2024). Previous research has investigated associations between late life depression and dementia (Blazer 2003, Kok 2017, Haigh 2018, Jacobs 2023, Kim 2021) or cognition (Shimada 2014, Wei 2019). However, since long-term cohorts are not available, previous research has been limited by self-reported life histories, which are prone to recall (Coughlin 1990) and survival biases (Hernan 2004 & 2017).

These difficulties exemplify common challenges of health research aiming to evaluate relationships between exposures and outcomes that occur many decades apart. For example, the lifecourse framework posits that exposures such as depression in early adulthood may influence mid- and late-life outcomes such as cognitive decline and dementia (Kuh 2003).

These influences are hypothesized to be mediated by decades-long social, behavioral, and pathological processes. Traditional cohorts would require decades of follow-up to investigate the association of prospectively-measured early-life exposures with late-life outcomes. However, few cohorts are available that include the comprehensive data from early through late life necessary to answer pressing questions in lifecourse epidemiology.

In the absence of an available cohort spanning the lifecourse, one way forward is to combine an early-life cohort (in which the exposure is measured) with a late-life cohort (in which the outcome is measured) to create a synthetic cohort with data on all life stages of interest. Using simple examples and simulations, we previously described the theoretical underpinnings for creating such a synthetic cohort using a matching approach (Kezios & Zimmerman 2024), based on a formal causal inference framework for pooling data sets - i.e. the “data fusion” problem (Barenboim 2016, Miao 2021, Breskin 2021, Cole 2022). Implementation of this method using real data sets requires that one evaluate whether pooling is reasonable given available cohorts, and to make several decisions about the matching process.

In this study, we apply this approach to estimate the effect of early adulthood depression on cognitive level and cognitive decline in mid-to late-life, employing two nationally representative cohorts from the United States: the 1979 National Longitudinal Survey of Youth (NLSY79; Bureau of Labor Statistics 2023) – spanning late adolescence to midlife – and the Health and Retirement Study (HRS) – spanning midlife to late life. These cohorts overlap in midlife, when participants are approximately 51-59 years old. We expand upon our previous theoretical and simulation results to detail the practical considerations that arise during implementation.

## Methods

### Overview of synthetic cohort construction

As detailed previously (Kezios & Zimmerman 2024), construction of a synthetic cohort from which inferences about associations between early life exposures and later-life outcomes can be validly estimated requires that several assumptions be met. Here we discuss decisions required at four stages -- (1) cohort selection, (2) data preparation, (3) linking data sets to generate a synthetic cohort using matching, and (4) statistical analysis and interpretation -- of synthetic cohort construction and analysis (Table 1).

**Table 1:** Synthetic cohort construction steps.

| Step | Description |
| --- | --- |
| 1. Cohort selection | Define the sample for which to make inferences based on the synthetic cohort. Identify necessary variables for identification based on a conceptual model. Evaluate whether 2 cohorts allow for identification of the parameter of interest based on the conceptual model and available data (e.g. drawn from the same underlying population, contain necessary variables that can be harmonized). |
| 2. Data preparation | Harmonize variables and handle missing data. |
| 3. Linking data sets to generate a synthetic cohort using matching | To create a synthetic cohort, fill in the structurally missing exposures and/or outcomes for participants in from your target cohort of interest by borrowing information from their matches in the other cohort, where matching is based on likely exposures given the confounders and mediators of the parameter of interest. |
| 4. Statistical analysis and interpretation | Analyze the synthetic cohort as you would a single cohort with both exposure and outcome measured, and interpret results in light of any limitations. Compute uncertainty measure accounting for multiple matches, if employed. |

### (1) Cohort selection

We aim to estimate the effect of earlier-life depression symptoms on later-life cognitive outcomes. In the absence of an available life course cohort, we aim to create a synthetic cohort spanning early adulthood through late life by combining an early-life cohort and later-life cohort. The first step is to choose an anchoring sample: the sample of participants about whom we aim to make inferences after we fill in information for the missing part of their life course. Here we have three options: 1) choose a later-life-sample as the anchor, and use the earlier-life sample to fill in the anchoring participants’ likely exposures (e.g., depression); 2) choose an earlier-life sample as the anchor, and use a later-life sample to fill in the anchoring participants’ likely outcomes (e.g., cognitive outcomes); and 3) choose the pooled earlier- and later-life samples as the anchor, and fill in likely outcomes and exposures, respectively.

Strategy 3 corresponds to a target estimand that is defined in the pooled samples, and is thus difficult to interpret, especially in the presence of selection processes that differ between the cohorts. Strategy 2 also involves an added complication of considering how to handle corrections for differential loss to follow-up. Thus, we focus on strategy 1, although strategy 2 would follow a similar process. The corresponding target estimand of interest is thus the effect of depression symptomatology in early adulthood on cognition in mid-to-late life among people who survive until mid-life and take part in the later-life study.

HRS is sponsored by the National Institute on Aging (grant number NIA U01AG009740) and is conducted by the University of Michigan. NLSY79 is sponsored and directed by the U.S. Bureau of Labor Statistics and managed by the Center for Human Resource Research (CHRR) at The Ohio State University. Interviews are conducted by the National Opinion Research Center (NORC) at the University of Chicago. The current analyses, using fully de-identified data, were not considered human subjects research.

#### Defining the later-life cohort of interest

We draw our later-life sample from HRS, an open cohort of participants aged 50+ and their spouses (who may have been less than 50 years old at enrollment) that began in 1992, and has since enrolled additional participants in five refresher waves for which data have been made available. Fielding of 10-item word recall tests began in 1998, prior to which 20-item word recall tests were fielded. Due to the potential for practice effects (i.e. improved test performance upon repeated exposures attributable to increasing familiarity with cognitive tests; Chen 2023, Wevue 2015, McCaffery 2000, Gross 2018), we limited our HRS sample of interest to all participants who were first interviewed and completed cognitive testing in 1998 or later (i.e. participants who were previously given a 20-item word recall test were excluded). Participants in this sample were reinterviewed up to eleven times, with interviews conducted approximately every other year between 2000 and 2020.

#### Selecting an earlier-life cohort from the same underlying population

The second step is to select another cohort whose participants may act as stand-ins to represent the early life experiences of the anchoring cohort. This is analogous to seeking exchangeable exposed and unexposed samples in conventional analyses. The most important consideration in this cohort selection is the plausibility that, conditional on measured covariates, stand-in cohort members represent the early life experiences of the anchoring cohort. This is plausible if participants in the two cohorts were drawn from the same underlying population. However, period effects and secular trends could threaten plausibility. Thus, when selecting a second cohort, it is important to assess the availability of measures obtained at similar ages for participants with similar birth years.

NLSY79 is a closed cohort that enrolled participants in adolescence/early adulthood, and has since followed them with annual or biannual interviews. Participants were aged 14-22 in 1979, and born between 1957 and 1965. Seventeen follow-up waves of data collected between 1982 and 2020 are available. Depression symptomatology was assessed beginning in 1992 (participants aged 27-35), and cognitive testing using 10-item word recall tests began in 2006 for participants aged 47+. Since all NLSY79 participants were over 47 years old at the most recent wave (participants were aged 55-63 in 2020), all NLSY79 participants who were administered word recall tests constitute potential stand-ins for the early-life experiences of the HRS participants.

A subset of HRS participants was born 1957-1964 and completed cognitive assessments when 47-63 years of age, similar to the NLSY79 cohort on both age and birth year (N=6,977 or 44% of participants): These participants were eligible to be part of synthetic cohort construction. In particular, the 2010 and later waves of HRS included substantial proportions of participants born 1957-1964 who were also in the age range 47-63 (30%-83% across waves). Other HRS participants completed their first cognitive assessments at age 47-63, similar to NLSY79 participants, but were not born between 1957 and 1965 (N=6,980 or 45% of participants).

Both cohorts were intended to be nationally representative of their respective target age groups at study enrollment, and the geographic components of the sampling frames for the two cohorts are similar: In NLSY79, households were drawn from non-overlapping primary sampling units across the US, with Hispanic participants oversampled. HRS includes households drawn from across the US in a multi-stage sampling design, with oversamples of Black participants, Hispanic participants, and residents of Florida (Schroeder 2023; Heeringa 1995). Based on US Census regions, the 2006 wave of NLSY includes South=42%, Midwest (i.e. “North Central”)=24%, West=19%, Northeast=15% (BLS 2025) HRS was similar in 2006: South=40%, Midwest=25%, West=19%, Northeast=16% (RAND 2025). Additionally, we are able to identify subsamples for whom midlife measures were obtained at approximately the same ages for participants born in the same time period, allowing us to limit the possibilities for cohort and period effects that differ between the two samples. Thus we have strong reasons to believe that we will be able to find pairs of similar participants that are drawn from the same underlying population.

#### Building a conceptual model

Exchangeable pairs of participants would have the same values of a set of covariates that d-separates the exposure and the outcome (i.e. block all front-door and back-door paths; Kezios and Zimmerman 2024). Finding such pairs of participants is theoretically possible so long as the positivity criterion is met: for all multivariate strata of the covariates that d-separate the exposure and outcome, participants in the early-life cohort must have a non-zero probability of being in the strata, so long as the corresponding probability is positive in the later-life cohort.

Multiple sets of covariates may grant d-separation. In order to determine possible sets, we start by drawing a conceptual causal directed acyclic graph (DAG): a representation of the hypothesized causal relationships between the exposure and outcome of interest. The DAG should detail all confounding and mediating paths, regardless of whether measures on each path are actually available in the cohorts of interest (Digitale 2021).

For our illustrative example, we identified a set of sociodemographic constructs likely to be confounders and a wide range of midlife health, sociodemographic, and behavioral constructs likely to be mediators of the relationship between depression in early adulthood and cognition in mid-to-late life (Figure 1). We include midlife versions of the exposure and the outcome (i.e. measures of midlife depression and midlife cognition) which, due to autocorrelation, are likely to be important mediators.

**Figure 1:**
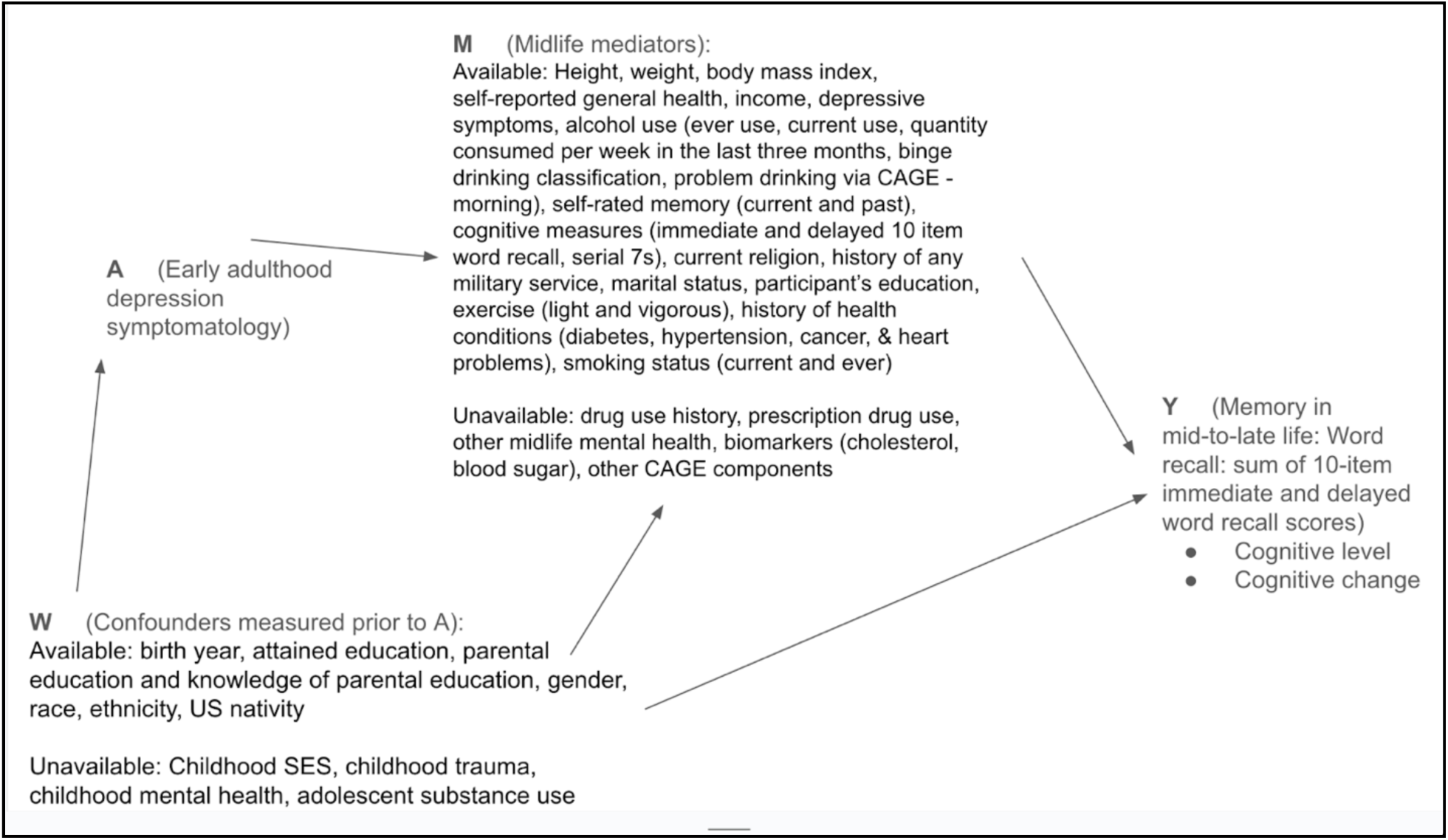
Causal DAG. We denote the confounders as **W** = (W_1,…W_p) and the mediators as **M** = (M_1,…M_q), and together these variables make up **X** (i.e. **X = W** ∪ **M)**. We denote the exposure A and the outcome Y.

#### Evaluating the available data

With a theory-based model of the relationships between the exposure and outcome in hand, we can now evaluate whether variables available in the candidate cohorts are sufficient to d-separate the exposure and the outcome in the causal DAG. These variables -- a subset of the confounders and mediators of the exposure-outcome association -- will be used to match individuals in the later-life anchoring cohort (for whom the outcome is measured) to individuals in the early-life matching cohort (for whom the exposure is measured). Additionally, since mediators evolve with age, potentially in a pattern that differs by birth cohort, we also matched on age and year of measurement.

#### Available Confounders

Although confounders in the DAG may include a wide range of covariates that could be measured prior to exposure in a hypothetical life course cohort, confounders available in both cohorts making up a synthetic cohort are likely to be limited to time-invariant variables, since variables for early-life constructs cannot be measured at the desired age for the later-life cohort. Thus, the quality and comprehensiveness of confounders in the later-life data set is likely to be inferior to the early-life data set. In some cases, the later-life cohort may include retrospective measures (e.g. “What was your income at the age of 18?” asked to participants in their 60s), however, these should be used cautiously, since differential recall by exposure or outcome can threaten validity (Coughlin 1990); the recalled version of the variable may not match the true early-life value of the variable if we were able to measure it for participants in the late-life cohort.

In our example, both NLSY79 and HRS included the following potential confounders, which we conceptualized as time-invariant as they were unlikely to change over time: birth year, gender, race, ethnicity, US nativity, knowledge of parental education, and, when known, respondent-reported parental education (as highest years of schooling). In NLSY79 we defined these variables using baseline data from 1979. In HRS, we used covariate values as recorded at enrollment.

#### Available Mediators

Mediators should be measured at similar ages in the two cohorts.

Both cohorts in our example included assessments of depressive symptoms (HRS-NLSY harmonized CES-D 6 point scale using the items for “Depressed”, “Effort”, “Restless”, “Lonely”, “Sad”, and “Going”, Appendix Table 5) and cognitive performance (immediate and delayed word recall separately, serial 7s score) completed during their overlapping age range (47-63 years). Other potential mediators measured in mid-life in both NLSY79 and HRS were: height, weight, body mass index, self-reported general health (1 = Excellent to 5 = Poor) household income (per-person, logged), alcohol use (ever use, current use, average number of drinks consumed per week in the last three months, frequency of drinking 6+ drinks in the last month (0 days, 1-3 days, 4-9 days,10 days or more)), CAGE “eye-opener” item), self-rated memory (current memory, change in memory; 1 = Excellent to 5 = Poor), current religion (Protestant, Catholic, Jewish, None, Other), history of any military service, marital status (Married, Separated/Divorced, Partnered, Widowed, Never married), participant’s education (Less than high school (HS), Some HS, HS, More than HS), exercise (light and vigorous separately, 1+ times/week), history of health conditions (diabetes, hypertension, cancer, & heart problems), smoking status (current and ever).

#### Assessing whether the set of available confounders and mediators is sufficient

Having identified the confounders and mediators in our DAG available in both cohorts, we now evaluate whether these available covariates are sufficient to d-separate the exposure and outcome (Figure 1). Among our pre-specified likely confounders, we were not able to include pre-baseline cognition, adverse childhood experiences or adolescent substance use. Among our pre-specified midlife mediators, we were not able to include factors such as prescription drug use and biomarker data. Although missing covariates along mediating pathways can attenuate findings in the synthetic cohort (Kezios & Zimmerman 2024), we believe that many of the mediators should be correlated (e.g. health behaviors and disease histories) and thus that it is unlikely that we are missing important covariates that would have independent causal relationships with the exposure and outcome (i.e. would sit on a path that could not be blocked by another mediator in our list).

### (2) Data Preparation

Having evaluated the available data, we can now prepare the data for matching.

#### Variable harmonization

The confounders and mediators identified as potential matching variables need to be harmonized between the cohorts. When questions are asked the same way in the two cohorts, and the range of potential answers is the same, no harmonization is necessary. In a slightly more complicated case, the questions are the same but the range of potential answers offered to participants differs. Categories may be combined in one of the data sets, or combined across multiple variables. For example, HRS coded race as three categories (“Black/African American”, “White/Caucasian” and “Other”), and two ethnicity categories (“Hispanic” and “Not Hispanic”). In contrast, NLSY79 asked for the race/ethnicity category with which the participant most strongly identified. Thus, in HRS we first created a race/ethnicity variable that prioritized coding participants as “Hispanic” if they endorsed Hispanic ethnicity, regardless of their endorsement of race. Because NLSY79 had more response options than HRS, we combined the NLSY79 responses to be congruent with the HRS race/ethnicity categories.

Differences in question wording can complicate harmonization efforts. In such cases, sensitivity analyses may be necessary to explore whether the results differ as a result of harmonizing differently. For example, the midlife versions of the items of the CES-D variable were asked on a 4-point scale in NLSY79 and a 2-point scale in HRS, and the wording in HRS was more vague. For our analysis, we prepared for sensitivity analyses by creating two harmonized versions of the 4-point NLSY79 scale, dichotomizing at thresholds of 1 and 2. We created a 6-point score consisting of the six items that could be harmonized between HRS and NLSY (Appendix Table 1).

#### Addressing missing data

A complete case approach leads to an undesirable reduction in sample size, and the severity of the sample size reduction is exacerbated as more variables are included in the matching covariate set. Thus imputation of missing data is desired to retain adequate sample size. However, since unbiased imputation of mediators and confounders requires knowledge of both the exposure and the outcome, we are unable to take advantage of imputation methods such as multiple imputation or structural equation models (MICE; Azur 2011, Lee 2010, Graham 2003, Schafer 1999). Further, because analyses in the synthetic cohort will have to account for multiple matches, simple methods for dealing with missing data are preferred at this stage. Such techniques include complete case analyses, conditional mean/median imputation, missing data indicators/categories, and carry/forward backward.

For each participant in each data set, we filled in missing values (HRS: 65% of initially missing observations, 4% of all observations; NLSY79: 33% of initially missing observations, 24% of all observations) by carrying forward the last observation.

### (3) Linking data sets to generate a synthetic cohort using matching

Once the two data sets are cleaned and harmonized, the analyst must decide how to link participants from the two cohorts. Following our previous theoretical work (Kezios & Zimmerman 2024), we aimed to match participants such that each later-life cohort member is matched to one or more earlier-life cohort members who reflect the early-life exposures of the later-life cohort member (i.e., are exchangeable). We achieve exchangeability by matching based on the set of available variables that best (given limitations imposed by available covariates) achieve d-separation the exposure and outcome and that were available in and harmonized between cohorts.

The properties of many matching algorithms have been extensively studied in real and simulated data (Greifer 2021, Cenzer 2020, Colson 2016, Ripollone 2020). Matching algorithms use one of several performance metrics to choose matches: they may aim to maximize the covariate balance across matched pairs, maximize propensity score (probability of belonging to one group or the other, given covariates) overlap, or minimize error between pairs’ covariate values in cross-validation. However, there is no single method that will optimize matching for the estimand and data of interest. Best practices prioritize optimizing covariate balance of all important covariates for the estimand of interest. Relying only on summary measures such as propensity score overlap can result in poor balance for individual matching variables.

Simulations are often the most effective strategy to choose the best approach (Colson 2016).

Our matching scheme is illustrated in Figures 2a-2c. We conducted coarsened exact matching on categorical variables to prioritize high-quality matches on these covariates. Baseline in HRS was defined as the first wave at which the participant’s cognition was measured. We required NLSY79 matches for a given HRS participant to be born within 5 years of that participant in the exact matching phase. Additionally, since both cohorts are longitudinal, we allowed any wave of NLSY79 to be a potential matching point for the HRS participants, but not vice versa, in line with our goal of filling in the likely early-life experiences of the HRS anchor cohort, by using exact matching on whether NLSY79 participants were within 5-years of the HRS participants’ age at each wave.

**Figure 2a:**
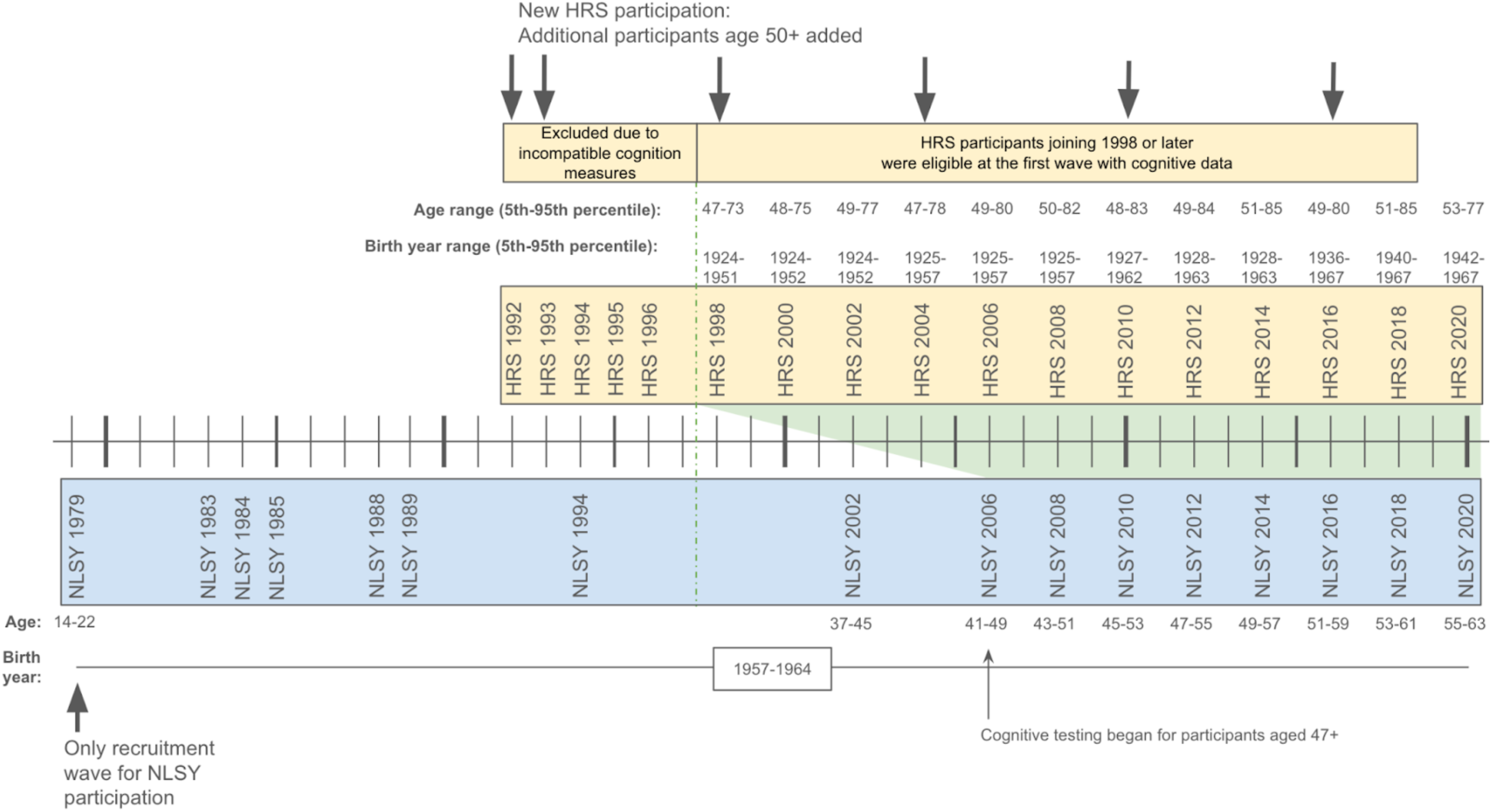
Matching Scheme - Cohort comparison. Graphical depiction comparing age, birth year, waves of data collection, and overlap in cognitive score measurement (green) in the Health and Retirement Study (HRS; yellow) to the National Longitudinal Study of Youth, 1979 (NLSY79; blue). HRS participants were recruited in 6 waves (top arrows) and followed longitudinally, and included respondents ages 51+ and their spouses of any age. NLSY participants were recruited in 1979 and followed longitudinally. In HRS, 10 item immediate and delayed word recall scores were ascertained. A 20 item scale was used before 1998, but this measure was incompatible with NLSY79 and thus participants enrolled before 1998 were excluded. Cognitive testing using the 10 item scale was included in NLSY79 starting at age 47 at the 2006 wave.

**Figure 2b:**
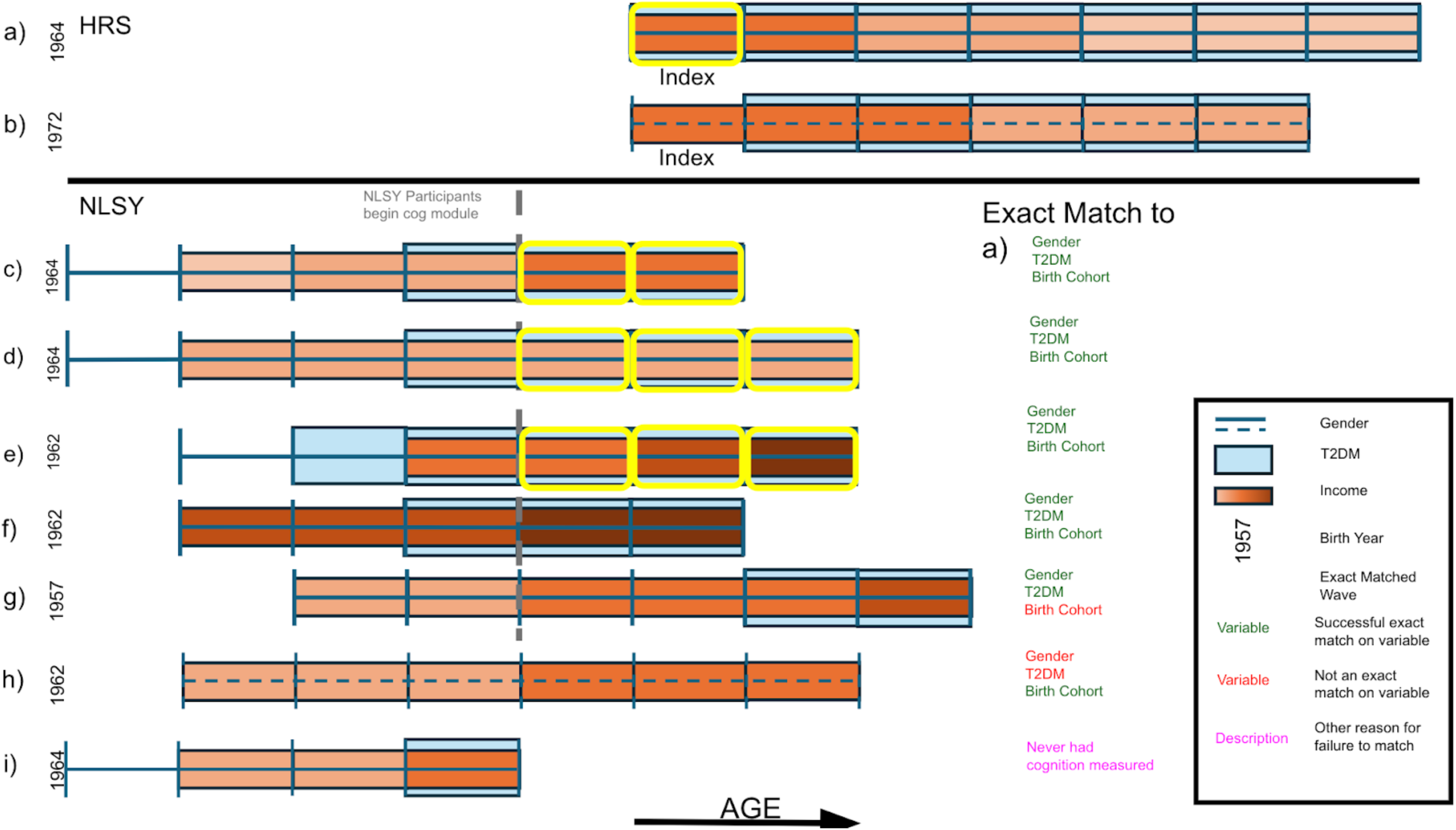
Matching Scheme - Exact Matching. Exact matching scheme for two hypothetical HRS participants: a) and b). Participant a)’s cognition was first measured at the index wave. They were followed for 7 waves. Participants c) – i) represent potential matches: hypothetical NLSY participants for whom we want to evaluate match quality with participant a)’s index wave. This hypothetical example demonstrates exact matching on gender identity, diabetes status (T2DM), and birth cohort (within 5 years of the index birth year): participants c) – f) exact matched participant a), and are eligible for the next step: distance matching. None of the NLSY participants were able to be exact matched to participant b) because no HRS participant was born within 5 years of their birth year.

**Figure 2c:**
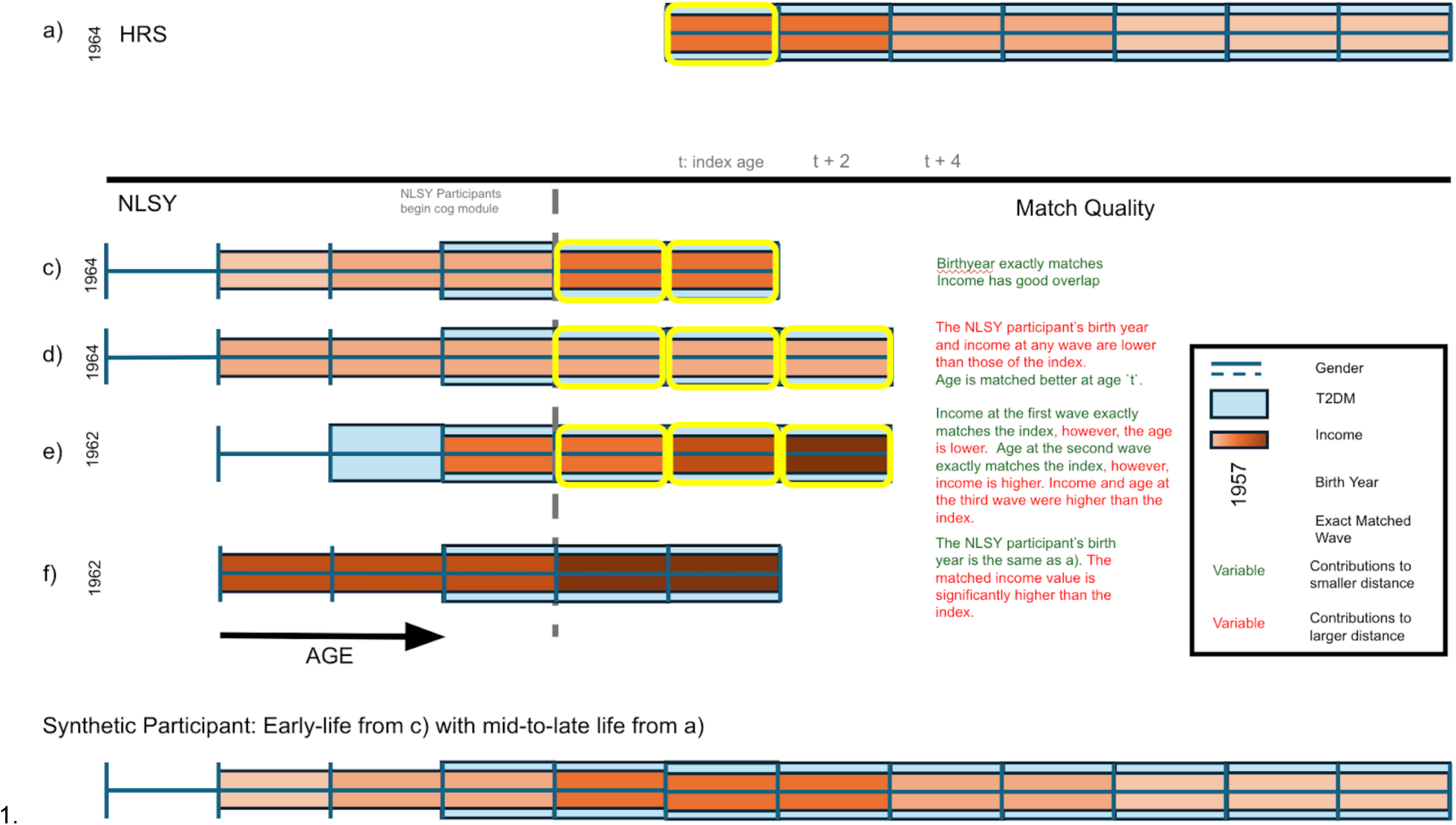
Matching Scheme - Distance Matching. Distance matching scheme for two hypothetical HRS participants. Participant a) was exact matched to participants c)-f) on gender identity, diabetes status (T2DM), and birth cohort (within 5 years of the index birth year). For each participant c)-f), we first aimed to find the wave in which the NLSY participant was most similar to the index. Participant c) had good overlap with participant a) in both eligible matching waves (t-2 and t), with similar income at each, however, the second matching (time t) wave was most similar in terms of age. Participant d) had similar income across all three eligible matching waves (t-2, t and t+2). The best matching-wave was the one that was closest in age (time t). For participant e), although income was a better match at the first matching wave (t-2) the second eligible matching wave (time t) was selected as the best matching wave as because age was identified as a higher priority co-variate compared to income. Participant f) had an income significantly higher than the index. This potential match was disqualified during match quality control. We next ranked the NLSY participants in terms of similarity with the index: Of the three remaining participants, Participant c) was the best match as they were most similar in terms of age and income. The bottom trajectory shows the example synthetic participant constructed form participants c) and a)

We used distance matching on continuous variables to further enhance exchangeability. Among the person-waves in NLSY79 matched with an HRS respondent, we selected matches with the lowest weighted Euclidean distance of standardized continuous variables. We standardized all continuous covariates by subtracting the mean and dividing by the standard deviation of the variable in HRS, as we wish to match NLSY79 participant-waves as closely as possible to the HRS participants – we did not use cohort-specific standardization because we did not want to match participants based on their relative position in each cohort.

We determined weights for the weighted Euclidean distances using an iterative approach that begins with a list of the continuous covariates. In each iteration, given the pool of remaining covariates, we calculated the sum of the two coefficients of determination (R^2^) from exposure-covariate and outcome-covariate models, conditional on the pool of previously included covariates in order to account for correlations among the matching variables. Thus in the first iteration we chose the most correlated covariate, in the second iteration we chose the next most correlated covariate, conditional on the first, and so on. This approach prioritized covariates that were most strongly associated with the exposure and/or outcome, while accounting for correlated variables. We calculated the marginal R^2^ as the difference in R^2^ between the first model including the variable and the model without the variable. We then created an associated weight as the inverse of the marginal R^2^ (Appendix 1)

Once distances were calculated, we selected the single best matching wave for the NLSY79 participant and selected up to 10 NLSY participants as matches for each HRS participant. We then randomly allocated the matched pairs to create 10 analytic datasets.

#### Optimizing the tradeoff between sample size and match quality

Depending on the hypothesized DAG, more than one subset of covariates may be sufficient for d-separation. Mediators may cause each other: for example, they may be chained together, such that controlling one of them may d-separate A from Y along the chain’s path. Thus the matching set need only include one of them. Likewise, for confounders, in some causal structures, controlling confounders downstream of other confounders. Matching covariates may be correlated in other ways such that matching on one variable will lead to an increase in the match quality of a variable not used in matching. Further, not all covariates are equally important, as some covariates have stronger relationships with the exposure and outcome.

An overarching consideration in choosing which set of covariates to match on (provided the set d-separates A from Y) is the trade-off between sample size and match quality: poor match quality leads to bias, while a smaller sample size increases noise. Matching on additional covariates generally reduces the quality of the match for previously-matched covariates, and exclusion of poor-quality matches reduces sample size. Since real-world samples are finite, differences in covariate values within a matched pair will occur. For example, despite the similarities in the HRS and NLSY79 discussed above, there may be important differences in the distributions of important covariates. In our case, although NLSY79 and HRS are likely to be drawn from the same population, there are small differences in the sampling schemes (e.g. oversampling of Black participants and Florida residents in HRS). Further, randomness, including in the selection of geographic sampling units, may lead to differences in the distributions of covariates in the two samples.

#### Prioritizing covariates

Due to the match quality vs smaller sample size tradeoff, it is not clear *a priori* which match set would create the best quality matches. Since we want to prioritize matching on the set of covariates that will lead to the best prediction of the exposure for each of the participants in the later-life cohort, variables that are strongly associated with both the exposure and the outcome are the most important variables to match on, as they account for a greater portion of the causal relationship linking the exposure and outcome. Thus, to optimize our analysis, we aim to choose a set of matching covariates that leads to good matches, and to prioritize high-quality matching for the variables that better explain the relationship between the exposure and outcome.

Variable importance methods quantify and rank variables in terms of how well they explain a dependent variable of interest. We again used the sum of R^2^ values from exposure-covariate and outcome-covariate models approach described above to define a priority order for matching. Thus we first matched on the variable (mediator or confounder) that explained the highest amount of the variability in the exposure-variable and outcome-variable, then matched on the variable that explained the highest amount of variability after accounting for the first variable, and so on.

The midlife versions of immediate recall, delayed recall, and CES-D score were among the most important variables, consistent with the intuition that these variables are autocorrelated within participants over time (Appendix Figure 1; Appendix Table 2). The alternative random forests approach yielded similar results (Appendix Figure 2; Appendix Table 3).

Another common algorithm for determining variable importance is based on ranking the mean decrease in predictive accuracy of an outcome (Nicodemus 2011, Jawadekar 2023) with removal of a given covariate in a random forest model. However, this approach does not account for the importance of a covariate conditional on other, more important, covariates. For comparison, we describe this alternate approach in Appendix 1.

#### Choosing the optimal set of matching covariates

Midlife CES-D was far more important than all other variables for explaining variation in early life CES-D, while midlife immediate recall, delayed recall, attained education, age and birth year explained later-life cognition, together explaining 15% of variation in the exposure and 42% of variation in the outcome. The contribution of additional variables was small (additional increase in sum of R^2 <0.01). Combining these two sets yielded our optimal matching set. The initial pre-matching samples included 7,751 NLSY participants and 20,813 HRS participants with cognitive measurements. After matching, the sample included 93,182 participant pairs (9,726 HRS participants and their corresponding 3,947 matched NLSY participants, mean matches per HRS participant: 9.58).

#### Evaluating match quality

To improve exchangeability, we impose match quality restrictions. In our final matched sample, we evaluated the match quality for each continuous matching covariate using a standardized difference measure defined as the difference in the covariate’s value between members of a pair, divided by the standard deviation of the covariate in the HRS sample. We dropped a matched pair if its standardized difference for any continuous covariate in the matching set was greater than a predefined cutoff, in terms of standard deviations of the covariate in HRS (moderate: 1.5SD, strict: 0.25SD).

In our applied example, sample sizes were reduced by imposing match quality cutoffs (1.5SD threshold: 78,153 matched pairs, n= 8,827 HRS participants and 4,911 unique matched NLSY participants, 0.25SD threshold: 28,060 matched pairs, n= 5,586 HRS participants and 3,947 matched NLSY participants).

### (4) Statistical analysis and Interpretation

#### Methods

Having selected an optimal synthetic cohort, we can finally analyze the data using our preferred analytic method as we would with a single-cohort analysis.

Briefly, we used confounder-adjusted linear mixed effects models with dichotomized 6-item CES-D as the exposure and sum of 10-item immediate and delayed word recall as the outcome. Separate models estimated cognitive level at age 50, cognitive level at age 65 and cognitive slope. We present estimates and 95% confidence intervals for coefficients pooled across mixed effects models. For the synthetic cohort, we estimated coefficients as the average coefficient across models estimated in the 10 analytic data sets, and we used Rubin’s rules (Rubin 1987) to estimate the variance of the coefficients. We adjusted for available time-invariant confounders (exposure, sex, US nativity, race/ethnicity, Armed Forces Qualification Test score (NLSY79 & synthetic only), education (participant’s & their parents’) and interview mode. We also adjusted for time-varying covariates at the wave prior to the exposure wave (NLSY79 & synthetic), or in the exposure wave (HRS), including religion, marital status, military service, & income. In HRS, we additionally adjusted for self-reported diabetes, hypertension, cancer, heart problems, physical activity, and general health.

We also estimated associations between CES-D and cognition in HRS and NLSY alone. Additionally, the analysis in NLSY79 acts as a test of the performance of the analysis of the synthetic cohort. Additional statistical methods details are provided in Appendix 2.

## Results

Prior to matching, using the 2006 wave of NLSY79 for comparison, NLSY79 had slightly higher average midlife CESD values (1.7 vs 1.4 on the 6-point scale), immediate (5.9 vs 5.7 points) and delayed (4.8 vs 4.6) word recall scores, and slightly lower attained education compared to HRS (Table 2). After matching, average values of the matching variables were very similar between HRS and NLSY79, but the values differed depending on which match quality threshold was imposed (Table 2). For example, the average CESD value with the 1.5 SD match quality threshold was 1.2 (for both HRS and NLSY79), while it was 1.4 with the 0.25 SD threshold.

**Table 2:** Sample characteristics for NLSY and HRS prior to matching. NLSY79 participants were eligible to be matched at seven waves. Missing NLSY79 values have been carried forward.

|  | NLSY sample of individuals eligible for matching in years exposure was measured |  | NLSY sample of individuals eligible for matching in years used to match to HRS participants for outcome data |  |  |  | HRS sample eligible for matching |
| --- | --- | --- | --- | --- | --- | --- | --- |
|  | 1983 | 1985 | 1988 | 1994 | 2002 | 2006 |  |
| Number of participants | 12221 | 10894 | 10465 | 8889 | 7723 | 7653 | 21263 |
| Height (inches) | 67.1 (4.1) | 67.1 (4.1) | 67.1 (4.1) | 67.1 (4.1) | 67.0 (4.1) | 67.2 (4.1) | 66.5 (4.1) |
| Height - Missing | 93 (0.8%) | 3 (0%) | 32 (0.3%) | 40 (0.4%) | 23 (0.3%) | 0 (0%) | 198 (0.9%) |
| Weight (lb) | 147.7 (30.3) | 152.8 (32.9) | 159.6 (35.9) | 170.8 (39.6) | 181.5 (43.0) | 185.9 (44.3) | 183.5 (44.5) |
| Weight - Missing | 95 (0.8%) | 3 (0%) | 0 (0%) | 0 (0%) | 0 (0%) | 0 (0%) | 271 (1.3%) |
| Body mass index | 23.0 (3.7) | 23.8 (4.2) | 24.8 (4.7) | 26.6 (5.5) | 28.3 (6.1) | 28.9 (6.1) | 29.1 (6.5) |
| Body mass index - Missing | 98 (0.8%) | 4 (0%) | 32 (0.3%) | 40 (0.4%) | 23 (0.3%) | 0 (0%) | 0 (0%) |
| General Health | Not Measured | Not Measured | Not Measured | Not Measured | 1.3 (1.0) | 1.4 (1.0) | 1.8 (1.2) |
| General Health - Missing | 12221 (100%) | 10894 (100%) | 10465 (100%) | 8889 (100%) | 2502 (32.4%) | 9 (0.1%) | 7 (0%) |
| Father's education (years) | 10.9 (3.8) | 10.8 (3.8) | 10.8 (3.8) | 10.8 (3.8) | 10.8 (3.8) | 10.8 (3.8) | 10.1 (4.4) |
| Father's education - Missing | 1721 (14.1%) | 1583 (14.5%) | 1518 (14.5%) | 1351 (15.2%) | 1140 (14.8%) | 1163 (15.2%) | 4626 (21.8%) |
| Mother's education (years) | 10.9 (3.1) | 10.8 (3.2) | 10.8 (3.2) | 10.8 (3.2) | 10.8 (3.2) | 10.8 (3.2) | 10.3 (4.1) |
| Mother's education - Missing | 757 (6.2%) | 700 (6.4%) | 667 (6.4%) | 590 (6.6%) | 490 (6.3%) | 498 (6.5%) | 2483 (11.7%) |
| Income (per person in household, log-transformed) | 1.2 (1.2) | 1.4 (1.3) | 1.6 (1.4) | 1.5 (1.3) | 1.6 (1.3) | 1.7 (1.4) | 2.1 (1.1) |
| Income - Missing | 11 (0.1%) | 2 (0%) | 0 (0%) | 0 (0%) | 0 (0%) | 0 (0%) | 0 (0%) |
| CES-D (6-point scale) | Not Measured | Not Measured | Not Measured | Not Measured | 1.7 (1.9) | 1.7 (1.9) | 1.4 (1.7) |
| CES-D - Missing | 12221 (100%) | 10894 (100%) | 10465 (100%) | 8889 (100%) | 5572 (72.1%) | 3027 (39.6%) | 1137 (5.3%) |
| Alcohol quantity (drinks/month) | Not Measured | Not Measured | 25.6 (45.1) | 23.3 (45.1) | 21.8 (38.0) | 20.7 (38.2) | 13.5 (33.4) |
| Alcohol quantity - Missing | 12221 (100%) | 10894 (100%) | 3494 (33.4%) | 1798 (20.2%) | 1343 (17.4%) | 1219 (15.9%) | 98 (0.5%) |
| Binge drinking | 0.6 (0.8) | 0.3 (0.7) | 0.5 (0.8) | 0.4 (0.8) | 0.2 (0.6) | 0.2 (0.7) | 0.3 (0.7) |
| Binge drinking - Missing | 7 (0.1%) | 0 (0%) | 0 (0%) | 1 (0%) | 0 (0%) | 1 (0%) | 5181 (24.4%) |
| **Self-rated memory (current)** | Not Measured | Not Measured | Not Measured | Not Measured | Not Measured | 2.6 (1.0) | 2.6 (1.0) |
| **Self-rated memory (current) - Missing** | 12221 (100%) | 10894 (100%) | 10465 (100%) | 8889 (100%) | 7723 (100%) | 6196 (81%) | 1031 (4.8%) |
| **Self-rated memory (past)** | Not Measured | Not Measured | Not Measured | Not Measured | Not Measured | 2.1 (0.5) | 2.1 (0.5) |
| **Self-rated memory (past) - Missing** | 12221 (100%) | 10894 (100%) | 10465 (100%) | 8889 (100%) | 7723 (100%) | 6196 (81%) | 1038 (4.9%) |
| **Immediate 10-word recall score** | Not Measured | Not Measured | Not Measured | Not Measured | Not Measured | 5.9 (1.8) | 5.7 (1.6) |
| **Immediate 10-word recall score - Missing** | 12221 (100%) | 10894 (100%) | 10465 (100%) | 8889 (100%) | 7723 (100%) | 6401 (83.6%) | 1161 (5.5%) |
| **Delayed 10-word recall score** | Not Measured | Not Measured | Not Measured | Not Measured | Not Measured | 4.8 (2.0) | 4.6 (1.9) |
| **Delayed 10-word recall score - Missing** | 12221 (100%) | 10894 (100%) | 10465 (100%) | 8889 (100%) | 7723 (100%) | 6538 (85.4%) | 1361 (6.4%) |
| **Serial 7s score** | Not Measured | Not Measured | Not Measured | Not Measured | Not Measured | 3.7 (1.5) | 3.6 (1.5) |
| **Serial 7s score - Missing** | 12221 (100%) | 10894 (100%) | 10465 (100%) | 8889 (100%) | 7723 (100%) | 6270 (81.9%) | 2156 (10.1%) |
| **Age at interview** | 22.3 (2.3) | 24.1 (2.2) | 27.6 (2.2) | 33.5 (2.2) | 41.4 (2.2) | 45.2 (2.2) | 55.4 (7.9) |
| **Age at interview - Missing** | 0 (0%) | 0 (0%) | 0 (0%) | 0 (0%) | 0 (0%) | 0 (0%) | 0 (0%) |
| **Birth year** | 1960.4 (2.2) | 1960.6 (2.2) | 1960.6 (2.2) | 1960.6 (2.2) | 1960.6 (2.2) | 1960.6 (2.2) | 1951.9 (12.0) |
| **Birth year - Missing** | 0 (0%) | 0 (0%) | 0 (0%) | 0 (0%) | 0 (0%) | 0 (0%) | 0 (0%) |
| **Current religion - Catholic** | 3796 (31.1%) | 3429 (31.5%) | 3254 (31.1%) | 2910 (32.7%) | 2035 (26.3%) | 2037 (26.6%) | 5602 (26.3%) |
| **Current religion - Other** | 1422 (11.6%) | 1235 (11.3%) | 1190 (11.4%) | 954 (10.7%) | 827 (10.7%) | 823 (10.8%) | 739 (3.5%) |
| **Current religion - None** | 6683 (54.7%) | 5939 (54.5%) | 5742 (54.9%) | 4812 (54.1%) | 4201 (54.4%) | 4157 (54.3%) | 2859 (13.4%) |
| **Current religion - Protestant** | 221 (1.8%) | 202 (1.9%) | 191 (1.8%) | 147 (1.7%) | 606 (7.8%) | 584 (7.6%) | 11703 (55%) |
| **Current religion - Jewish** | 97 (0.8%) | 87 (0.8%) | 86 (0.8%) | 64 (0.7%) | 54 (0.7%) | 52 (0.7%) | 286 (1.3%) |
| **Current religion - Missing** | 2 (0%) | 2 (0%) | 2 (0%) | 2 (0%) | 0 (0%) | 0 (0%) | 74 (0.3%) |
| **Ever drank alcohol** | 11424 (93.5%) | 10298 (94.5%) | 9974 (95.3%) | 7217 (81.2%) | 6302 (81.6%) | 6223 (81.3%) | 18449 (86.8%) |
| **Ever drank alcohol - Missing** | 5 (0%) | 0 (0%) | 0 (0%) | 1 (0%) | 5 (0.1%) | 7 (0.1%) | 0 (0%) |
| **Currently drink alcohol** | 8069 (66%) | 7215 (66.2%) | 6996 (66.9%) | 5319 (59.8%) | 4150 (53.7%) | 3953 (51.7%) | 14093 (66.3%) |
| **Currently drink alcohol - Missing** | 6 (0%) | 0 (0%) | 0 (0%) | 1 (0%) | 0 (0%) | 1 (0%) | 6 (0%) |
| **Military service** | 2071 (16.9%) | 1166 (10.7%) | 1157 (11.1%) | 1065 (12%) | 915 (11.8%) | 919 (12%) | 3164 (14.9%) |
| **Military service - Missing** | 0 (0%) | 0 (0%) | 0 (0%) | 0 (0%) | 0 (0%) | 0 (0%) | 7 (0%) |
| **Marital status - Married** | 7241 (59.3%) | 5402 (49.6%) | 3521 (33.6%) | 1923 (21.6%) | 1439 (18.6%) | 1351 (17.7%) | 15205 (71.5%) |
| **Marital status - separated/divorced/absent** | 4386 (35.9%) | 4740 (43.5%) | 5832 (55.7%) | 5618 (63.2%) | 4530 (58.7%) | 4374 (57.2%) | 3505 (16.5%) |
| **Marital status - never married** | 582 (4.8%) | 739 (6.8%) | 1085 (10.4%) | 1307 (14.7%) | 1682 (21.8%) | 1820 (23.8%) | 1460 (6.9%) |
| **Marital status - widowed** | 12 (0.1%) | 13 (0.1%) | 27 (0.3%) | 41 (0.5%) | 72 (0.9%) | 108 (1.4%) | 1072 (5%) |
| **Marital status - Missing** | 0 (0%) | 0 (0%) | 0 (0%) | 0 (0%) | 0 (0%) | 0 (0%) | 21 (0.1%) |
| **Education** | 1.7 (0.7) | 1.8 (0.7) | 1.9 (0.7) | 1.9 (0.7) | 1.9 (0.7) | 1.9 (0.7) | 2.0 (0.8) |
| **Education - Missing** | 39 (0.3%) | 33 (0.3%) | 31 (0.3%) | 21 (0.2%) | 18 (0.2%) | 21 (0.3%) | 121 (0.6%) |
| **Light exercise** | 0 (0%) | 0 (0%) | 0 (0%) | 0 (0%) | 5605 (72.6%) | 5475 (71.5%) | 11721 (55.1%) |
| **Light exercise - Missing** | 12221 (100%) | 10894 (100%) | 10465 (100%) | 8889 (100%) | 140 (1.8%) | 251 (3.3%) | 5587 (26.3%) |
| **Vigorous exercise** | 0 (0%) | 0 (0%) | 0 (0%) | 0 (0%) | 2882 (37.3%) | 2809 (36.7%) | 7165 (33.7%) |
| **Vigorous exercise - Missing** | 12221 (100%) | 10894 (100%) | 10465 (100%) | 8889 (100%) | 142 (1.8%) | 253 (3.3%) | 34 (0.2%) |
| **Diabetes** | 0 (0%) | 0 (0%) | 0 (0%) | 0 (0%) | 283 (3.7%) | 410 (5.4%) | 3499 (16.5%) |
| **Diabetes - Missing** | 12221 (100%) | 10894 (100%) | 10465 (100%) | 8889 (100%) | 2505 (32.4%) | 12 (0.2%) | 0 (0%) |
| **Hypertension** | 0 (0%) | 0 (0%) | 0 (0%) | 0 (0%) | 901 (11.7%) | 1321 (17.3%) | 9181 (43.2%) |
| **Hypertension - Missing** | 12221 (100%) | 10894 (100%) | 10465 (100%) | 8889 (100%) | 2505 (32.4%) | 12 (0.2%) | 0 (0%) |
| **Cancer** | 0 (0%) | 0 (0%) | 0 (0%) | 0 (0%) | 100 (1.3%) | 155 (2%) | 1525 (7.2%) |
| **Cancer - Missing** | 12221 (100%) | 10894 (100%) | 10465 (100%) | 8889 (100%) | 2507 (32.5%) | 11 (0.1%) | 0 (0%) |
| **Heart problems** | 0 (0%) | 0 (0%) | 0 (0%) | 0 (0%) | 146 (1.9%) | 212 (2.8%) | 2735 (12.9%) |
| **Heart problems - Missing** | 12221 (100%) | 10894 (100%) | 10465 (100%) | 8889 (100%) | 2507 (32.5%) | 11 (0.1%) | 0 (0%) |
| **Ever smoker** | 0 (0%) | 0 (0%) | 0 (0%) | 0 (0%) | 0 (0%) | 0 (0%) | 11853 (55.7%) |
| **Ever smoker - Missing** | 12221 (100%) | 10894 (100%) | 10465 (100%) | 8889 (100%) | 7723 (100%) | 7653 (100%) | 7 (0%) |
| **Current smoker** | 0 (0%) | 0 (0%) | 0 (0%) | 0 (0%) | 0 (0%) | 0 (0%) | 4936 (23.2%) |
| **Current smoker - Missing** | 12221 (100%) | 10894 (100%) | 10465 (100%) | 8889 (100%) | 7723 (100%) | 7653 (100%) | 11 (0.1%) |
| **CAGE score** | 0 (0%) | 185 (1.7%) | 186 (1.8%) | 158 (1.8%) | 138 (1.8%) | 145 (1.9%) | 1330 (6.3%) |
| **CAGE score - Missing** | 12221 (100%) | 2486 (22.8%) | 1736 (16.6%) | 1535 (17.3%) | 1304 (16.9%) | 1291 (16.9%) | 3653 (17.2%) |

In NLSY79, where we have early adulthood CES-D and midlife cognitive measures, a one point higher CES-D in early adulthood was associated with a −0.023 (95%CI:-0.032, −0.015) point difference in average summed recall during follow up (“cognitive level”), but no difference in words recalled per CES-D point per decade of follow-up (“cognitive slope”; 0.004 (95%CI:-0.034, 0.042) points per decade) (Table 4, Figure 3a).

**Figure 3a:**
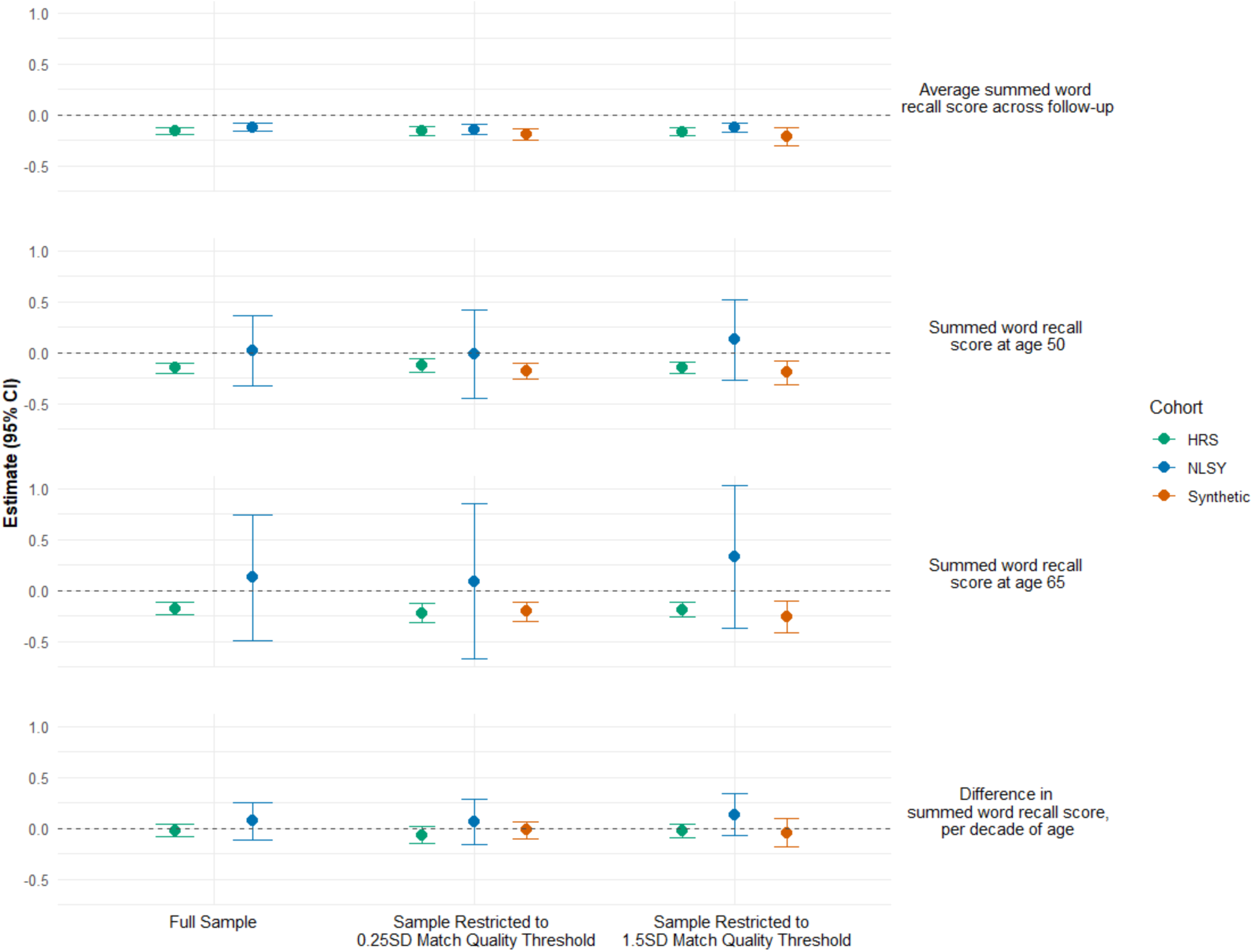
Estimated associations of depressive symptomatology age 50-56 in NLSY, HRS and combined synthetic cohorts with cognitive level and change capped at age 63. To improve harmonization, follow-up ages in HRS and the synthetic cohorts were limited to ages 63 and younger to match NLSY’s available follow-up, and exposures in the NLSY and synthetic cohorts were from NLSY’s 50s module to match HRS’s exposure assessment. Covariate-adjusted associations of depressive symptomatology (6-item CES-D) with average summed word recall (A) was estimated in a model without an age slope term. Separate models were used to estimate associations with summed word recall scores at ages 50 and 65 (intercepts; B & C), but the estimated associations with difference in summed word recall per decade of age (slopes; D) from the two models were identical. Both match quality thresholds were for the match set that used midlife CES-D, midlife immediate word recall, midlife delayed word recall, attained education, age and birth year as matching covariates, and, for the HRS and NLSY cohorts, restricted to participants who were part of at least one matched pair in the synthetic cohort.

**Table 3:** Sample characteristics of matched individuals in the synthetic cohorts in the year of the match, comparing narrow (0.25 SD) versus wide (1.5 SD) match quality calipers.

| Match quality threshold | 0.25 SD |  | 1.5 SD |  |
| --- | --- | --- | --- | --- |
| Cohort | NLSY | HRS | NLSY | HRS |
| Height (inches) | 67.3 (4.1) | 66.8 (4.0) | 67.0 (4.2) | 66.6 (4.1) |
| Height - Missing | 0 (0%) | 62 (0.2%) | 0 (0%) | 413 (0.5%) |
| Weight (lb) | 186.7 (44.4) | 193.5 (46.7) | 184.7 (44.6) | 190.3 (46.5) |
| Weight - Missing | 0 (0%) | 231 (0.8%) | 0 (0%) | 862 (1%) |
| Body mass index | 28.9 (6.0) | 30.4 (6.8) | 28.8 (6.1) | 30.1 (6.8) |
| Body mass index - Missing | 0 (0%) | 0 (0%) | 0 (0%) | 0 (0%) |
| General Health | 1.4 (1.0) | 1.7 (1.1) | 1.4 (1.0) | 1.8 (1.1) |
| General Health - Missing | 3 (0%) | 4 (0%) | 9 (0%) | 16 (0%) |
| Father's education (years) | 11.5 (3.6) | 11.4 (3.7) | 11.1 (3.9) | 10.9 (4.1) |
| Father's education - Missing | 1880 (6.3%) | 1880 (6.3%) | 10423 (12%) | 10423 (12%) |
| Mother's education (years) | 11.4 (3.0) | 11.7 (3.1) | 11.0 (3.4) | 11.1 (3.6) |
| Mother's education - Missing | 336 (1.1%) | 336 (1.1%) | 2994 (3.5%) | 2994 (3.5%) |
| Income (per person in household, log-transformed) | 1.9 (1.4) | 2.2 (1.2) | 1.8 (1.5) | 2.1 (1.1) |
| Income - Missing | 0 (0%) | 0 (0%) | 0 (0%) | 0 (0%) |
| CES-D (6-point scale) | 1.2 (1.5) | 1.2 (1.5) | 1.4 (1.6) | 1.4 (1.6) |
| CES-D - Missing | 0 (0%) | 0 (0%) | 0 (0%) | 0 (0%) |
| Alcohol quantity (drinks/month) | 18.2 (31.6) | 15.8 (33.3) | 18.2 (33.3) | 15.2 (34.1) |
| Alcohol quantity - Missing | 3647 (12.2%) | 52 (0.2%) | 11271 (13%) | 252 (0.3%) |
| **Binge drinking** | 0.2 (0.6) | 0.4 (0.7) | 0.2 (0.6) | 0.4 (0.7) |
| **Binge drinking - Missing** | 0 (0%) | 6757 (22.6%) | 0 (0%) | 20731 (23.9%) |
| **Self-rated memory (current)** | 2.5 (0.9) | 2.5 (0.9) | 2.5 (1.0) | 2.6 (1.0) |
| **Self-rated memory (current) - Missing** | 12 (0%) | 3 (0%) | 62 (0.1%) | 13 (0%) |
| **Self-rated memory (past)** | 2.1 (0.4) | 2.1 (0.5) | 2.1 (0.5) | 2.1 (0.5) |
| **Self-rated memory (past) - Missing** | 1 (0%) | 18 (0.1%) | 12 (0%) | 72 (0.1%) |
| **Immediate 10-word recall score** | 6.0 (1.2) | 6.0 (1.2) | 5.9 (1.5) | 5.9 (1.4) |
| **Immediate 10-word recall score - Missing** | 0 (0%) | 0 (0%) | 0 (0%) | 0 (0%) |
| **Delayed 10-word recall score** | 5.0 (1.5) | 5.0 (1.5) | 4.8 (1.6) | 4.8 (1.7) |
| **Delayed 10-word recall score - Missing** | 0 (0%) | 0 (0%) | 0 (0%) | 0 (0%) |
| **Serial 7's score** | 3.9 (1.4) | 3.8 (1.4) | 3.8 (1.4) | 3.7 (1.5) |
| **Serial 7's score - Missing** | 483 (1.6%) | 384 (1.3%) | 2260 (2.6%) | 2037 (2.3%) |
| **Age at interview** | 53.0 (2.4) | 53.0 (2.3) | 52.7 (2.8) | 52.6 (3.1) |
| **Age at interview - Missing** | 0 (0%) | 0 (0%) | 0 (0%) | 0 (0%) |
| **Birth year** | 1960.3 (2.5) | 1960.1 (2.9) | 1960.0 (2.6) | 1959.1 (4.1) |
| **Birth year - Missing** | 0 (0%) | 0 (0%) | 0 (0%) | 0 (0%) |
| **Current religion - Catholic** | 7467 (25%) | 6409 (21.5%) | 21945 (25.3%) | 20292 (23.4%) |
| **Current religion - Other** | 2124 (7.1%) | 1726 (5.8%) | 6735 (7.8%) | 4654 (5.4%) |
| **Current religion - None** | 3124 (10.5%) | 5137 (17.2%) | 9095 (10.5%) | 14315 (16.5%) |
| **Current religion - Protestant** | 16935 (56.7%) | 16147 (54.1%) | 48347 (55.7%) | 46244 (53.3%) |
| **Current religion - Jewish** | 210 (0.7%) | 357 (1.2%) | 616 (0.7%) | 997 (1.1%) |
| **Current religion - Missing** | 0 (0%) | 84 (0.3%) | 0 (0%) | 236 (0.3%) |
| **Ever drank alcohol** | 24754 (82.9%) | 27782 (93%) | 71239 (82.1%) | 79233 (91.3%) |
| **Ever drank alcohol - Missing** | 19 (0.1%) | 0 (0%) | 109 (0.1%) | 0 (0%) |
| **Currently drink alcohol** | 17828 (59.7%) | 22411 (75.1%) | 49742 (57.3%) | 62962 (72.6%) |
| **Currently drink alcohol - Missing** | 0 (0%) | 1 (0%) | 0 (0%) | 10 (0%) |
| **Military service** | 3333 (11.2%) | 3198 (10.7%) | 9154 (10.6%) | 8475 (9.8%) |
| **Military service - Missing** | 0 (0%) | 6 (0%) | 0 (0%) | 20 (0%) |
| **Marital status - Married** | 3481 (11.7%) | 3120 (10.4%) | 11173 (12.9%) | 8548 (9.9%) |
| **Marital status - separated/divorced/absent** | 18503 (62%) | 20023 (67.1%) | 51028 (58.8%) | 59484 (68.6%) |
| **Marital status - never married** | 7193 (24.1%) | 5951 (19.9%) | 22405 (25.8%) | 16520 (19%) |
| **Marital status - widowed** | 683 (2.3%) | 747 (2.5%) | 2132 (2.5%) | 2142 (2.5%) |
| **Marital status - Missing** | 0 (0%) | 19 (0.1%) | 0 (0%) | 44 (0.1%) |
| **Education** | 2.2 (0.5) | 2.2 (0.5) | 2.1 (0.7) | 2.1 (0.7) |
| **Education - Missing** | 0 (0%) | 0 (0%) | 0 (0%) | 0 (0%) |
| **Light exercise** | 22558 (75.5%) | 23579 (79%) | 64011 (73.8%) | 66282 (76.4%) |
| **Light exercise - Missing** | 968 (3.2%) | 1 (0%) | 3730 (4.3%) | 664 (0.8%) |
| **Vigorous exercise** | 11597 (38.8%) | 9169 (30.7%) | 32254 (37.2%) | 25715 (29.6%) |
| **Vigorous exercise - Missing** | 970 (3.2%) | 26 (0.1%) | 3748 (4.3%) | 101 (0.1%) |
| **Diabetes** | 3518 (11.8%) | 4596 (15.4%) | 10391 (12%) | 14331 (16.5%) |
| **Diabetes - Missing** | 0 (0%) | 0 (0%) | 8 (0%) | 0 (0%) |
| **Hypertension** | 9346 (31.3%) | 13000 (43.5%) | 26622 (30.7%) | 37766 (43.5%) |
| **Hypertension - Missing** | 0 (0%) | 0 (0%) | 6 (0%) | 0 (0%) |
| **Cancer** | 834 (2.8%) | 1896 (6.3%) | 2538 (2.9%) | 5645 (6.5%) |
| **Cancer - Missing** | 0 (0%) | 0 (0%) | 0 (0%) | 0 (0%) |
| **Heart problems** | 1367 (4.6%) | 3001 (10.1%) | 3960 (4.6%) | 9063 (10.4%) |
| **Heart problems - Missing** | 6 (0%) | 0 (0%) | 26 (0%) | 0 (0%) |
| **Ever smoker** | 878 (2.9%) | 15952 (53.4%) | 2797 (3.2%) | 47023 (54.2%) |
| **Ever smoker - Missing** | 15396 (51.6%) | 0 (0%) | 45520 (52.5%) | 3 (0%) |
| **Current smoker** | 5822 (19.5%) | 7103 (23.8%) | 17439 (20.1%) | 21981 (25.3%) |
| **Current smoker - Missing** | 1045 (3.5%) | 0 (0%) | 3294 (3.8%) | 8 (0%) |
| **CAGE score** | 363 (1.2%) | 1925 (6.4%) | 1216 (1.4%) | 6097 (7%) |
| **CAGE score - Missing** | 4866 (16.3%) | 2094 (7%) | 15745 (18.2%) | 7576 (8.7%) |

Time varying characteristics refer to the value for the year/wave the NLSY and HRS participants were matched (e.g., 1988, 1994, 2002, or 2006 for NLSY).

**Table 4:** Associations of depressive symptom score in early adulthood with mid-to-late life cognition in NLSY79.

| Term | Coefficient (95% CI) [SE] |  |  |  |  |  |
| --- | --- | --- | --- | --- | --- | --- |
|  | Harmonized results using all available follow-up & CESD measured at age 50-56 |  |  | Results using all available follow-up & CESD in 1992 at age 27-35 |  |  |
|  | NLSY cognition regressed on NLSY CESD in the full NLSY sample | NLSY cognition regressed on NLSY CESD among NLSY participants included in the matched synthetic cohorts |  | NLSY cognition regressed on NLSY in the full NLSY sample | NLSY cognition regressed on NLSY CESD among NLSY participants included in the matched synthetic cohorts |  |
|  |  | 1.5 SD Match Quality Threshold | 0.25 SD Match Quality Threshold |  | 1.5 SD Match Quality Threshold | 0.25 SD Match Quality Threshold |
| Model without interaction |  |  |  |  |  |  |
| Average summed word recall score across follow-up | -0.120 (-0.161, -0.078) [0.021] | -0.124 (-0.171, -0.076) [0.024] | -0.140 (-0.192, -0.089) [0.026] | -0.023 (-0.032, -0.015) [0.005] | -0.026 (-0.036, -0.016) [0.005] | -0.023 (-0.034, -0.013) [0.005] |
| Models with interaction |  |  |  |  |  |  |
| Summed word recall score at age 50 | 0.018 (-0.325, 0.361) [0.175] | 0.131 (-0.264, 0.526) [0.202] | -0.010 (-0.441, 0.421) [0.221] | -0.016 (-0.087, 0.055) [0.036] | -0.015 (-0.097, 0.067) [0.042] | 0.028 (-0.060, 0.116) [0.045] |
| Summed word recall score at age 65 | 0.127 (-0.485, 0.740) [0.313] | 0.330 (-0.370, 1.030) [0.358] | 0.091 (-0.672, 0.855) [0.391] | -0.010 (-0.137, 0.118) [0.065] | -0.006 (-0.151, 0.140) [0.074] | 0.069 (-0.087, 0.225) [0.080] |
| Difference in summed word recall score, per decade of age | 0.073 (-0.108, 0.254) [0.092] | 0.133 (-0.072, 0.337) [0.104] | 0.068 (-0.155, 0.291) [0.114] | 0.004 (-0.034, 0.042) [0.019] | 0.006 (-0.037, 0.048) [0.022] | 0.027 (-0.019, 0.072) [0.023] |

Models adjusted for CES-D, sex, US nativity, race/ethnicity, Armed Forces Qualification Test score, education (participant's & their parents') and interview mode, religion, marital status, military service, & income.

In HRS, where we have midlife CES-D and mid-to-late life cognitive measures, a one point higher CES-D in midlife was associated with a −0.163 (95%CI:-0.199,-0.128) point difference in average cognitive level but no difference in cognitive slope (−0.021 (95%CI:-0.062,0.020) points per decade) (Table 5, Figure 3b).

**Figure 3b:**
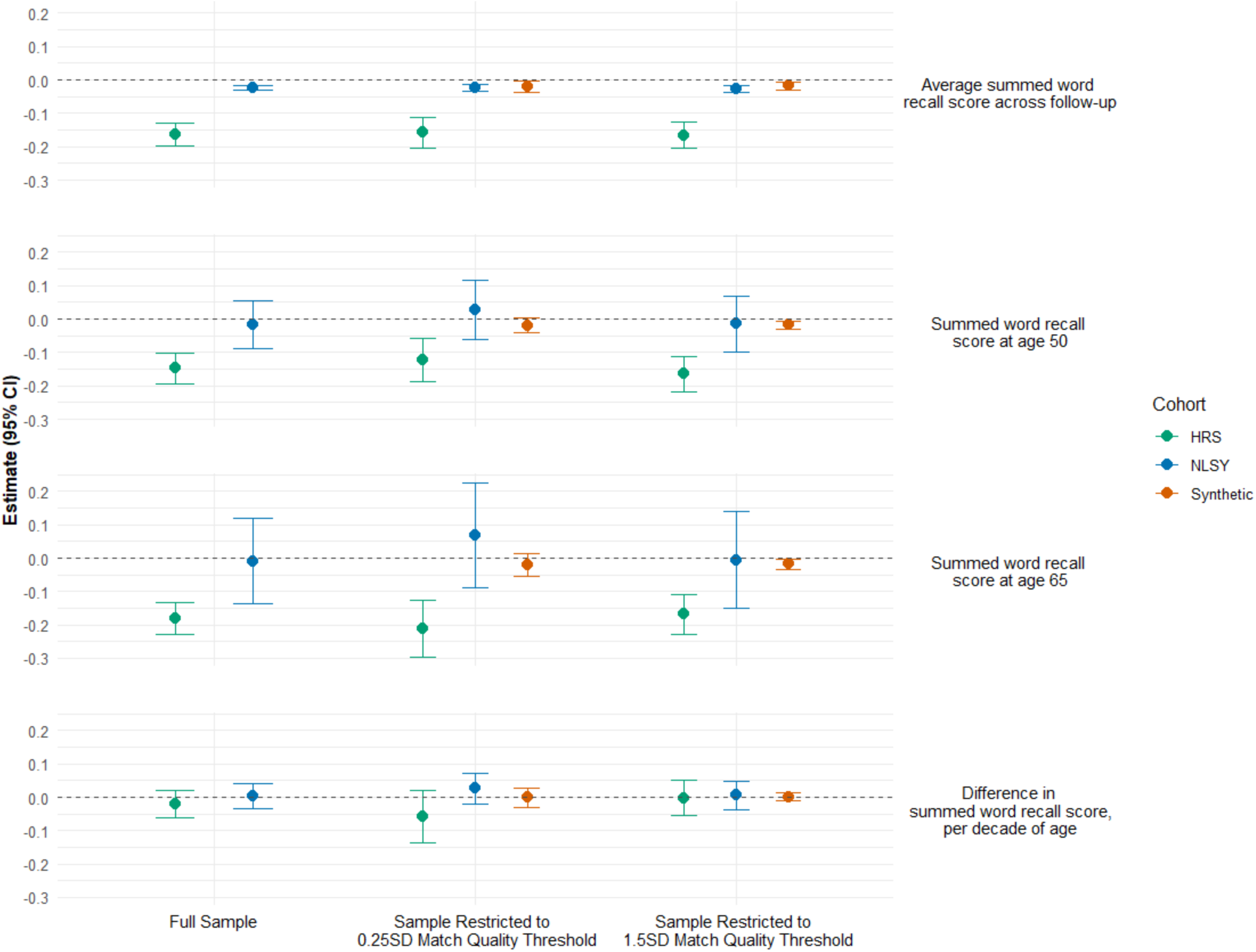
Estimated associations of earliest available depressive symptom scores in NLSY, HRS and combined synthetic cohorts with cognitive level and change, using all available follow-up. Extended results that relax harmonization restrictions to include earliest available exposure and all available follow-up for HRS and the synthetic cohort. All available follow-up was used for all three cohorts: thus NLSY’s maximum follow-up age was 63, while HRS and the synthetic cohort had a maximum follow-up age of 79. Exposure (CES-D) was measured at ages 50-56 in HRS, and ages 27-35 for NLSY and the synthetic cohort. Covariate-adjusted associations of depressive symptomatology (6-item CES-D, with each item dichotomized) with average summed word recall (A) was estimated in a model without an age slope term. Separate models were used to estimate associations with summed word recall scores at ages 50 and 65 (intercepts; B & C), but the estimated associations with difference in summed word recall per decade of age (slopes; D) from the two models were identical. Both match quality thresholds were for the match set that used midlife CES-D, midlife immediate word recall, midlife delayed word recall, attained education, age and birth year as matching covariates, and, for the HRS and NLSY cohorts, restricted to participants who were part of at least one matched pair in the synthetic cohort.

**Table 5:** Associations of depressive symptom score in midlife with mid-to-late life cognition in HRS.

| Term | Coefficient (95% CI) [SE] |  |  |  |  |  |
| --- | --- | --- | --- | --- | --- | --- |
|  | Harmonized results restricting maximum age to 63, CESD measured at age 50-56 |  |  | Results using all available follow-up & CESD measured at age 50-56 |  |  |
|  | HRS Cognition regressed on HRS CES-D in the full, harmonized HRS sample | HRS cognition regressed on HRS CES-D among HRS participants included in the matched synthetic cohorts |  | HRS Cognition regressed on HRS CES-D in the full HRS sample | HRS cognition regressed on HRS CES-D among HRS participants included in the matched synthetic cohorts |  |
|  |  | 1.5 SD Match Quality Threshold | 0.25 SD Match Quality Threshold |  | 1.5 SD Match Quality Threshold | 0.25 SD Match Quality Threshold |
| Model without interaction |  |  |  |  |  |  |
| Average summed word recall score across follow-up | -0.159 (-0.195, -0.124) [0.018] | -0.163 (-0.202, -0.124) [0.02] | -0.157 (-0.203, -0.111) [0.023] | -0.163 (-0.199, -0.128) [0.018] | -0.166 (-0.206, -0.127) [0.02] | -0.158 (-0.204, -0.112) [0.023] |
| Models with interaction |  |  |  |  |  |  |
| Summed word recall score at age 50 | -0.147 (-0.197, -0.097) [0.026] | -0.148 (-0.203, -0.093) [0.028] | -0.119 (-0.185, -0.052) [0.034] | -0.148 (-0.194, -0.103) [0.023] | -0.165 (-0.217, -0.113) [0.027] | -0.123 (-0.189, -0.058) [0.033] |
| Summed word recall score at age 65 | -0.176 (-0.238, -0.115) [0.031] | -0.186 (-0.257, -0.116) [0.036] | -0.219 (-0.309, -0.128) [0.046] | -0.180 (-0.228, -0.132) [0.024] | -0.168 (-0.229, -0.108) [0.031] | -0.212 (-0.298, -0.127) [0.044] |
| Difference in summed word recall score, per decade of age | -0.020 (-0.077, 0.037) [0.029] | -0.026 (-0.090, 0.039) [0.033] | -0.067 (-0.151, 0.017) [0.043] | -0.021 (-0.062, 0.020) [0.021] | -0.002 (-0.056, 0.051) [0.027] | -0.059 (-0.138, 0.020) [0.040] |

ES-D measures were on a 6 point scale, where each point represents the presence of a dichotomized depression symptom. Models adjusted for sex, US nativity, race/ethnicity, education (participant's & their parents'), interview mode, religion, marital status, military service, income, self-reported diabetes, hypertension, cancer, heart problems, physical activity, and general health.

When limiting to similar ages of exposure measurement (50-56) and restricting maximum age of follow-up (age 63) that overlapped in HRS and NLSY79, a one point higher CES-D was associated with lower average cognitive level in both NLSY79 (−0.120 (95%CI:-0.161,-0.078)) and HRS (−0.159 (95%CI:-0.195,-0.124)), and both thresholds for the synthetic cohort (1.5SD: - 0.216 (95%CI:-0.306,-0.125); 0.25SD: −0.188 (95%CI:-0.244,-0.132)). For cognitive slope, we observed no association for NLSY79 (0.073 (95%CI:-0.108,0.254)), HRS (−0.020 (95%CI:-0.077,0.037)), or the synthetic cohort (1.5SD: −0.043 (95%CI:-0.182,0.095); 0.25SD: −0.016 (95%CI:-0.103,0.070)). Wide confidence intervals for models estimating cognitive slopes in NLSY79 illustrate the limited number of follow-up cognitive measurements in this age range. Results were similar when restricting to participants included in the matched synthetic cohorts at both match thresholds. (Tables 4-6, Figure 3a).

**Table 6:** Associations of early adulthood depression scores with mid-to-late life cognition in synthetic cohorts created by combining NLSY79 and HRS.

| Term | Coefficient (95% CI) [SE] |  |  |  |
| --- | --- | --- | --- | --- |
|  | HRS cognition regressed on NLSY79 CES-D among matched pairs |  |  |  |
|  | Harmonized results restricting maximum age to 63 & using CESD measured at age 50-56 |  | Results using all available follow-up & CESD in 1992 at age 27-35 |  |
|  | 1.5 SD Match Quality Threshold | 0.25 SD Match Quality Threshold | 1.5 SD Match Quality Threshold | 0.25 SD Match Quality Threshold |
| **Model without interaction** |  |  |  |  |
| **Average summed word recall score across follow-up** | -0.216 (-0.306, -0.125)<br>[0.046] | -0.188 (-0.244, -0.132)<br>[0.029] | -0.018 (-0.030, -0.006)<br>[0.006] | -0.020 (-0.037, -0.003)<br>[0.009] |
| **Models with interaction** |  |  |  |  |
| **Summed word recall score at age 50** | -0.193 (-0.312, -0.074)<br>[0.061] | -0.179 (-0.256, -0.103)<br>[0.039] | -0.018 (-0.032, -0.005)<br>[0.007] | -0.019 (-0.040, 0.002)<br>[0.011] |
| **Summed word recall score at age 65** | -0.258 (-0.417, -0.099)<br>[0.081] | -0.204 (-0.299, -0.110)<br>[0.048] | -0.017 (-0.033, -0.002)<br>[0.008] | -0.021 (-0.055, 0.013)<br>[0.017] |
| **Difference in summed word recall score, per decade of age** | -0.043 (-0.182, 0.095)<br>[0.071] | -0.016 (-0.103, 0.070)<br>[0.044] | 0.001 (-0.011, 0.012)<br>[0.006] | -0.001 (-0.029, 0.027)<br>[0.014] |

Exposure was the NLSY79 participants' early adulthood CES-D. Each of the six items comprising the CES-D score was dichotomized at $\geq 1$ . The coefficients refer to expected difference in outcomes per one point increment in this score (range: 0-6). Cognition was from the HRS participant's first measured cognition wave starting at age 48. Models adjusted for sex, US nativity, race/ethnicity, Armed Forces Qualification Test score, education (participant's & their parents') and interview mode, religion, marital status, military service, & income.

Similarity in the harmonized results supports interpretation of the non-harmonized results in the synthetic cohort, where we have mid-to-late life cognition (from HRS participants) and early adulthood CES-D (from their matched NLSY79 participants). When using a moderate cutoff for match quality (1.5SD), a one-point increase in CES-D in early adulthood was associated with a lower cognitive level (−0.018 (95%CI:-0.030,-0.006) point difference), but no difference in cognitive slope (0.001 (95%CI:-0.011,0.012) points per decade) in mid-to-late life. When using a stricter cutoff, match quality improved and sample size decreased, but estimates were substantively similar (0.25SD threshold: −0.020 (95%CI:-0.037,-0.003) point difference in cognitive level and −0.001 (95%CI:-0.029,0.027) points per decade difference in cognitive slope; Table 6).

## Discussion

Combining evidence from two large, nationally representative cohorts, we found that depressive symptoms in early adulthood predict memory level in middle to older ages, but have no clear association with rate of memory decline. For comparisons in which the analysis using the synthetic cohort could be compared directly to analyses using single cohorts, findings were very similar.

Extensive research links depression and cognitive outcomes, including dementia (Blazer 2003, Kok 2017, Haigh 2018, Jacobs 2023, Kim 2021, Shimada 2014, Wei 2019). Our findings are consistent with this, but do not provide evidence that depressive symptoms predict faster rate of memory loss over time. A major challenge in this literature -- as with much of lifecourse epidemiology -- is the paucity of data sets spanning the entire lifecourse. We demonstrate an approach based on synthetic data generated by combining information from early- and late-life cohorts. Analyzing synthetic cohorts created by pooling existing data sources can generate useful new knowledge of life course exposure and disease patterns that we would otherwise be unable to observe without fielding new long-term studies or waiting for current cohorts to reach advanced age.

However, like all analytic approaches, creating a synthetic cohort has potential pitfalls, which fall into two categories. First, to achieve exchangeability, it is important that participants be drawn from the same underlying population, but this is often not verifiable. Samples may come from different underlying populations for unknown reasons. In our example analysis, although both samples are nationally-representative, we may question the extent to which the two samples contain participant pairs who, conditional on observed covariates, can be considered exchangeable. Cohort effects pose a particular challenge: pooling by exactly matching on birth year and age limits our follow-up. Using less strict matching requirements on birth year is appealing, allowing for the creation of synthetic cohorts with longer follow-up, the introduced mismatch in birth year may reduce our confidence in the result, as the life course experiences of matched pairs of individuals no longer line up with true chronological time. For example, early-life data from an individual born in 1960 and thus age 40 in 2000 could be matched at age 40 to late-life data from a similar individual born in 1940 and age 40 in 1980. However, the generated synthetic life course would re-use calendar years 1980-2000, during which myriad contextual factors may impact the exposure-outcome relationship. Second, researchers must decide on a list of variables to match on, a decision we recommend guiding by drawing a causal DAG, but there is always the possibility that the DAG does not contain all true causal pathways due to limited prior knowledge. The DAG clarifies the variables required to d-separate the exposure and outcome, which should be matched on, but not all matching variables of interest may be available in both data sets. These variables are likely correlated, and thus the effect of omitted variables may be small if they are not strong confounders or mediators. Matching on a variable also leads to improved quality of matches on the variables correlated with the matching variable, even if they are not observed. We do not specifically describe the relationships among mediators and confounders in our causal structure. However, if strong information is available it could be used to better identify minimal subsets of matching variables.

In our example, results were aligned between the synthetic cohort and the results in NLSY79 alone, suggesting that we were not missing important mediators and did not have substantial unmeasured confounding, and were able to find high-quality matches. However this increase in match quality came at the cost of reduced sample size, decreasing the number of HRS participants by about 9% for the 1.5SD quality control threshold and 42% for the 0.25SD threshold.

Our framework illustrates how pooling data to generate a synthetic cohort involves making many decisions that may affect synthetic cohort validity. Some decisions are straightforward: for example variables may need to be harmonized to be comparable across datasets. Others are more difficult: Choosing a set of variables to match on is plagued by a trade-off between sample size and match quality. Dropping poor-quality matches may modify the estimand if match quality is correlated with covariates in either cohort. For example, if participants with cancer history are rare, it will be more difficult to find high-quality matches for those participants, and they may be disproportionately dropped. The estimand of interest may thus need to be modified, or selection weights used to account for this sample modification.

At each step we highlighted an approach we took, but there are many other possible analytic paths one could take. These methodological developments provide a promising direction for combining information across multiple cohorts, and future improvements may facilitate implementation. One promising direction is to leverage a third “crosswalking” data set in which both versions of questions are asked to participants in order to improve harmonization.

Additionally, methods based on weighting or multi-sample g-computation (Robins 1986) rather than matching may have different challenges, such as the need for correct model specification, but may result in larger sample sizes, and should be explored in future work.

### Conclusion

In the absence of available traditional cohorts, data pooling approaches have emerged as a way forward to estimating life course effects on late life outcomes. Using matching to combine data from two or more data sources spanning separate, but overlapping, parts of the life course is one such promising approach, but it requires careful implementation to ensure that assumptions are met and estimates are appropriately interpreted.

## Data Availability

All data produced in the present study are publicly available.

## Appendix

### Appendix 1 Alternate method for prioritizing covariates – Sum of R-squareds

To illustrate, we fit separate random forest models predicting the exposure from midlife covariates and time-invariant confounders in NLSY79, and the outcome from the same covariates in HRS. In NLSY79 we used midlife covariates from midlife to predict CES-D from 1992. In HRS we used midlife covariates from the first wave at which cognition was measured to predict later-life cognition among the participants with at least 4 waves of follow-up; 42% of the HRS sample participated in at least 4 waves, and 28% of those were at least 65 by wave 4 (mean=62.4, median=60).

For both the exposure and the outcome, random forest tuning indicated that out-of-bag (OOB) error was negligibly different across cutoffs of number of variables per split, so we used the default value (8). OOB error was stable when using 500 trees.

Note how this strategy is the same as the way we created the weights for distance matching. We quantified variable importance by iteratively estimating the sum of the two coefficients of determination (R^2^) from exposure-covariate and outcome-covariate models, conditional on the pool of previously chosen covariates in order to account for correlations among the matching variables. In each iteration, we chose the variable corresponding to the highest R^2^ to include in future models, and calculated the marginal R^2^ as the difference in R^2^ between the model including the variable and the model without the variable. We created an associated weight as the inverse of the marginal R^2^. This approach prioritized covariates that were most strongly associated with the exposure and/or outcome, while accounting for correlated variables, and resulted in both an ordering of the variables from most to least important, and associated weights. We then constructed a series of matching sets, starting with only the most important covariate, and iteratively adding each next important covariate to each subsequent matching set. We repeated the matching process described above on each matching set, resulting in a set of synthetic cohorts.

Associations of each matching variable with the exposure and outcomes, and the resulting order in which we iteratively added covariates to the matching set, are presented in Appendix Table 2 and Appendix Figure 1. Since the 20 point scale is the gold standard exposure, we used the variable ordering from the 20 point scale to determine all match sets.

### Appendix 2 Statistical analysis method details

#### Exposure: depression scores based on CES-D items

Depressive symptoms were quantified using the items from the Center for Epidemiologic Studies Depression score. We chose the 1992 NLSY79 wave, when participants were aged 27-35 to define our exposure. This 1992 wave was the first time the CES-D items were administered. For each of the 20 items making up the CES-D questionnaire, respondents were asked to assess the past-week frequency of each symptom in the CES-D, in the form of statements such as “During the past week, I was *bothered by things that usually don’t bother me*” (Never; one to two days/week; three to four days/week; five to seven days/week). Each item was scored from 0 (never) to 3 (five to seven days/week) and the scores for all items were summed for a final CES-D score. All symptoms assessed in the CES-D are presented in Appendix Table 1.

HRS assessed six of the CES-D items reported in NLSY, and they were asked as binary variables (“Much of the time during the past week, you felt _ “).

Thus we created a 6-point score (CES-D-6) consisting of the six items that could be harmonized between HRS and NLSY (Appendix Table 1) We then created dichotomized versions of the NLSY79 variables dichotomized at >= 1 (CES-D.1, CES-D-SF.1, CES-D-6.1) and >=2 (CES-D.2, CES-D-SF.2, CES-D-6.2). While our exposure of interest was the 20-item CES-D scale measured in NLSY79, these versions better allow for comparison with estimated associations of earlier-life depression symptoms and mid-life cognition in NLSY79, or mid-life depression and later-life cognition in HRS. To put these measures on the same scale, we divided by the number of items and maximum value of the item.

#### Outcome: cognitive score based on word recall

NLSY cognitive assessments – fielded starting in 2006 at the first interview when participants were at least age 47 – were selected by NLSY to mirror the assessments in HRS. In both surveys, word recall assessments were conducted by reading the participant a list of 10 words, and asking the participant to immediately recall as many of the 10 words as they could (“immediate recall test”). Then, after a delay of a few minutes, participants were again asked to recall as many of the 10 words as possible (“delayed recall test”). For both the immediate and delayed recall tests, we created scores for each participant of the number of correctly-recalled words from a list of 10, and then added these two scores together to create a summed recall score.

We used word recall scores from 2006-2016 in NLSY79 as potential mediators. In HRS, ten-word immediate and delayed word recall tests were administered from 1996 to 2020. We used covariates from the first wave at which cognition was measured in HRS as mediators.

Cognitive assessments are known to improve with repeated exposure, a phenomenon referred to as “practice effects,” and typically addressed by adjusting cognitive scores based on extent of prior exposure to the test. In preliminary analysis in HRS, cognitive test performance reached a maximum at the third attempt, on average. To account for these differences in performance due to previous exposure to cognitive test materials, we created a variable for the count of times the participant had taken the test (“test count”). We then modeled summed recall in a linear mixed effects model using this count and all covariates from the adjustment set, except CES-D. Preliminary work showed that cognitive test performance peaked during the third attempt. We included the practice effect from the third attempt as an offset for the first attempt, and the difference between the third and second attempts as an offset for the second attempt. The interpretation of coefficients is thus the association between covariates and cognition after participants become accustomed to the cognitive testing procedure. We additionally included an indicator for interview mode (web/phone or in person).

Using the coefficient on the test count in the third test exposure as an offset, we estimated associations of CES-D during midlife with cognitive level at age 50 and cognitive change thereafter using linear mixed-effects model adjusted for time-invariant covariates.

In order to maximize power and avoid reverse correlation between coefficients for cognitive level and cognitive slope, we ran separate models to estimate cognitive level at age 50 and cognitive slope. We estimate average cognitive level conditional on covariates using the following model form, which assumes that CESD has same effect regardless of age at cognitive measurement:

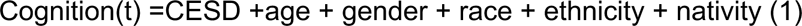

We the allowed for interaction between CESD and age in order to estimate cognitive slope:

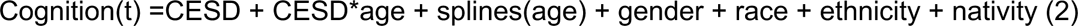

We interpret the coefficients on CESD in model (1) as the associations of depressive score in a given year with cognitive level at age 50, and the coefficients on the interaction terms in model (2) as the change in cognition per decade of age for a one point increase in CESD. Note that the appropriate set of adjustment covariates for our parameters of interest includes pre-exposure covariates defined by our time-invariant confounders, and including other matching covariates would amount to controlling for mediators. While it may be tempting to include other pre-exposure covariates from the NLSY79 participant in each matched pair, there is no guarantee that these covariates would accurately reflect the early life experiences of the HRS participants unless the matching procedure also matched on all confounders and mediators between the adjustment variable and the outcome. As this is unlikely, we do not include other adjustment variables in the adjustment set.

We present estimates and 95% confidence intervals for coefficients pooled across mixed effects models. We estimated coefficients as the average coefficient across models estimated in the 10 data sets, and we used Rubin’s rules for pooling estimates in order to estimate the variance in the estimate. Our primary parameters of interest were the association of CESD with cognition at age 50 and on cognitive decline, as measured by the interaction term for CESD and age. For comparison we also present the age slope in cognitive score, representing average cognitive decline per decade of age.

Additionally, we conducted linear regression of the association of earlier-life depression scores with mid-life cognition NLSY79 alone, and mixed-effects modeling of mid-life depression with mid-to-late-life cgnition in HRS alone using the same method as in the synthetic cohort.

**Appendix Table 1:** CES-D component items and exposure variable comparisons.

| Full CES-D item | CES-D-SF | CES-D-6 |
| --- | --- | --- |
| I WAS BOTHERED BY THINGS THAT USUALLY DON'T BOTHER ME... |  |  |
| I DID NOT FEEL LIKE EATING; MY APPETITE WAS POOR... | X |  |
| I FELT THAT I COULDN'T SHAKE OFF THE BLUES EVEN WITH HELP FROM MY FAMILY AND FRIENDS... |  |  |
| I FELT THAT I WAS JUST AS GOOD AS OTHER PEOPLE... |  |  |
| I HAD TROUBLE KEEPING MY MIND ON WHAT I WAS DOING... |  |  |
| I FELT DEPRESSED... | X | X |
| I FELT THAT EVERYTHING I DID WAS AN EFFORT... | X | X |
| I FELT HOPEFUL ABOUT THE FUTURE... |  |  |
| I THOUGHT MY LIFE HAD BEEN A FAILURE... |  |  |
| I FELT FEARFUL... |  |  |
| MY SLEEP WAS RESTLESS... | X | X |
| I WAS HAPPY... |  |  |
| I TALKED LESS THAN USUAL... |  |  |
| I FELT LONELY... | X | X |
| PEOPLE WERE UNFRIENDLY... |  |  |
| I ENJOYED LIFE... |  |  |
| I HAD CRYING SPELLS... |  |  |
| I FELT SAD... | X | X |
| I FELT THAT PEOPLE DISLIKE ME... |  |  |
| I COULD NOT GET "GOING"... | X | X |

**Appendix Table 2:** Associations of each matching variable with the exposure and outcome R-squared based.

| Matching Variable | Association with exposure (R^2) | Association with outcome (R^2) | Sum of R^2s for associations with exposure and outcome | Incremental increase in R^2 when adding variable to the multivariate exposure-covariate and outcome-covariate models |
| --- | --- | --- | --- | --- |
| IMMED_RECALL | 0.0196 | 0.3391 | 0.3587 | 0.3587 |
| CESD_NEW6PT | 0.1256 | 0.3424 | 0.468 | 0.1092 |
| DELAY_RECALL | 0.1258 | 0.3923 | 0.5181 | 0.0502 |
| EDU_NEW | 0.1473 | 0.4040 | 0.5513 | 0.0331 |
| BIRTHYEAR | 0.1488 | 0.4178 | 0.5666 | 0.0154 |
| MARRIAGE | 0.1558 | 0.4180 | 0.5738 | 0.0072 |
| GENHEALTH_ORD | 0.1598 | 0.4195 | 0.5793 | 0.0055 |
| CAGE_MORNING | 0.1644 | 0.4198 | 0.5842 | 0.0049 |
| SERIAL_7 | 0.1686 | 0.4213 | 0.5899 | 0.0057 |
| HEIGHT | 0.1718 | 0.4225 | 0.5943 | 0.0044 |
| SMOKE_EVER | 0.1748 | 0.4228 | 0.5976 | 0.0033 |
| ALCOHOL_DRINKSPERMO | 0.1773 | 0.4229 | 0.6002 | 0.0026 |
| HYPERTENSION | 0.1805 | 0.4233 | 0.6038 | 0.0036 |
| ALCOHOL_NOW | 0.1828 | 0.4235 | 0.6063 | 0.0024 |
| SMOKE_NOW | 0.1846 | 0.4237 | 0.6083 | 0.0020 |
| MILITARY | 0.1855 | 0.4245 | 0.61 | 0.0017 |
| RELIGIONCURRENT | 0.1870 | 0.4245 | 0.6115 | 0.0016 |
| RATE_MEM_COG | 0.1879 | 0.4251 | 0.613 | 0.0014 |
| ALCOHOL_EVER | 0.1892 | 0.4251 | 0.6143 | 0.0013 |
| RATE_PAST_COG | 0.1900 | 0.4252 | 0.6152 | 0.0010 |
| DAD_EDU | 0.1903 | 0.4257 | 0.616 | 0.0008 |
| VIG_EXERCISE | 0.1912 | 0.4257 | 0.6169 | 0.0010 |
| DIABETES | 0.1917 | 0.4260 | 0.6177 | 0.0007 |
| MOM_EDU | 0.1920 | 0.4262 | 0.6182 | 0.0005 |
| BMI | 0.1925 | 0.4262 | 0.6187 | 0.0005 |
| LIGHT_EXERCISE | 0.1925 | 0.4266 | 0.6191 | 0.0003 |
| ALCOHOL_BINGE2_ORD | 0.1928 | 0.4266 | 0.6194 | 0.0003 |
| WEIGHT | 0.1929 | 0.4266 | 0.6195 | 0.0001 |
| INCOME_PP_LOG10 | 0.1929 | 0.4265 | 0.6194 | 0.0001 |
| HEARTPROB | 0.1929 | 0.4266 | 0.6195 | 0.0000 |
| CANCER | 0.1929 | 0.4266 | 0.6195 | 0.0000 |

**Appendix Table 3:** Random forest variable importance results.

| Midlife covariate | NLSY (mean decrease in accuracy for exposure model) | HRS (mean decrease in accuracy for outcome model) | Sum of variable importance associations with exposure and outcome |
| --- | --- | --- | --- |
| CES-D (6 point) | 41.86 | 11.43 | 53.29 |
| Immediate recall | 3.5 | 48.51 | 52.01 |
| Education | 12.09 | 39.09 | 51.18 |
| Delayed recall | 2.01 | 43.51 | 45.52 |
| Age | 5.88 | 37.16 | 43.04 |
| Birth year | 6.86 | 34.63 | 41.49 |
| Weight | 13.16 | 24.55 | 37.71 |
| BMI | 13.71 | 21.16 | 34.87 |
| Race/ethnicity | 10.41 | 20.93 | 31.34 |
| General health | 8.24 | 19.38 | 27.62 |
| Height | 8.78 | 18 | 26.78 |
| Gender | 8.28 | 17.64 | 25.92 |
| Mother's attained education | 5.76 | 15.47 | 21.23 |
| Serial 7s score | 6.99 | 12.96 | 19.95 |
| Father's education | 5.02 | 14.56 | 19.58 |
| Marital status | 10.45 | 9.04 | 19.49 |
| Income per person (log 10) | 4.04 | 14.6 | 18.64 |
| Knowledge of mother's attained education | 2.44 | 11.67 | 14.11 |
| Smoking: ever | 7.8 | 5.12 | 12.92 |
| Smoking: current | 4.74 | 7.59 | 12.33 |
| Self-rated current memory | -0.06 | 12.27 | 12.21 |
| Knowledge of father's attained education | 3.04 | 8.82 | 11.86 |
| History of diabetes | 1.9 | 9.35 | 11.25 |
| Military history | 1.59 | 9.55 | 11.14 |
| Light exercise | 4.2 | 5.91 | 10.11 |
| Hypertension | 1.16 | 8.14 | 9.3 |
| US Birth | -0.18 | 9.18 | 9 |
| Alcohol<br>(Drinks/month) | 4.14 | 4.2 | 8.34 |
| Current religion | 1.32 | 4.29 | 5.61 |
| Alcohol (CAGE<br>morning) | 2.67 | 2.79 | 5.46 |
| Alcohol (Binge<br>drinking) | 0.24 | 5.06 | 5.3 |
| Vigorous exercise | 0.2 | 4.84 | 5.04 |
| Alcohol (ever drinking) | -0.14 | 4.9 | 4.76 |
| History of heart<br>problems | -0.07 | 4.53 | 4.46 |
| Alcohol (current<br>drinking) | 2.27 | 2.11 | 4.38 |
| Self-rated past<br>memory | 4.41 | -0.41 | 4 |
| History of cancer | -1.35 | -0.86 | -2.21 |

**Appendix Figure 1:**
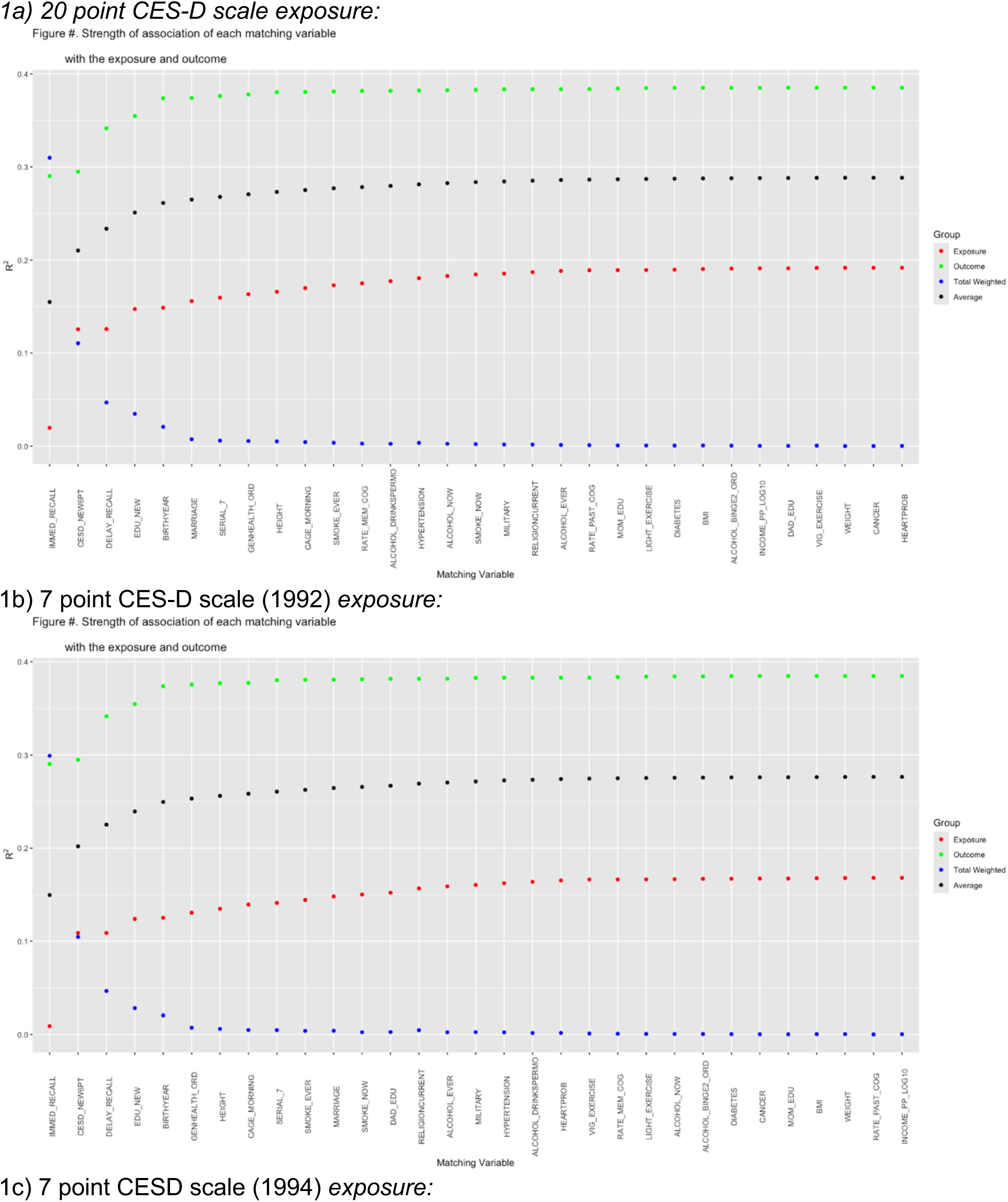
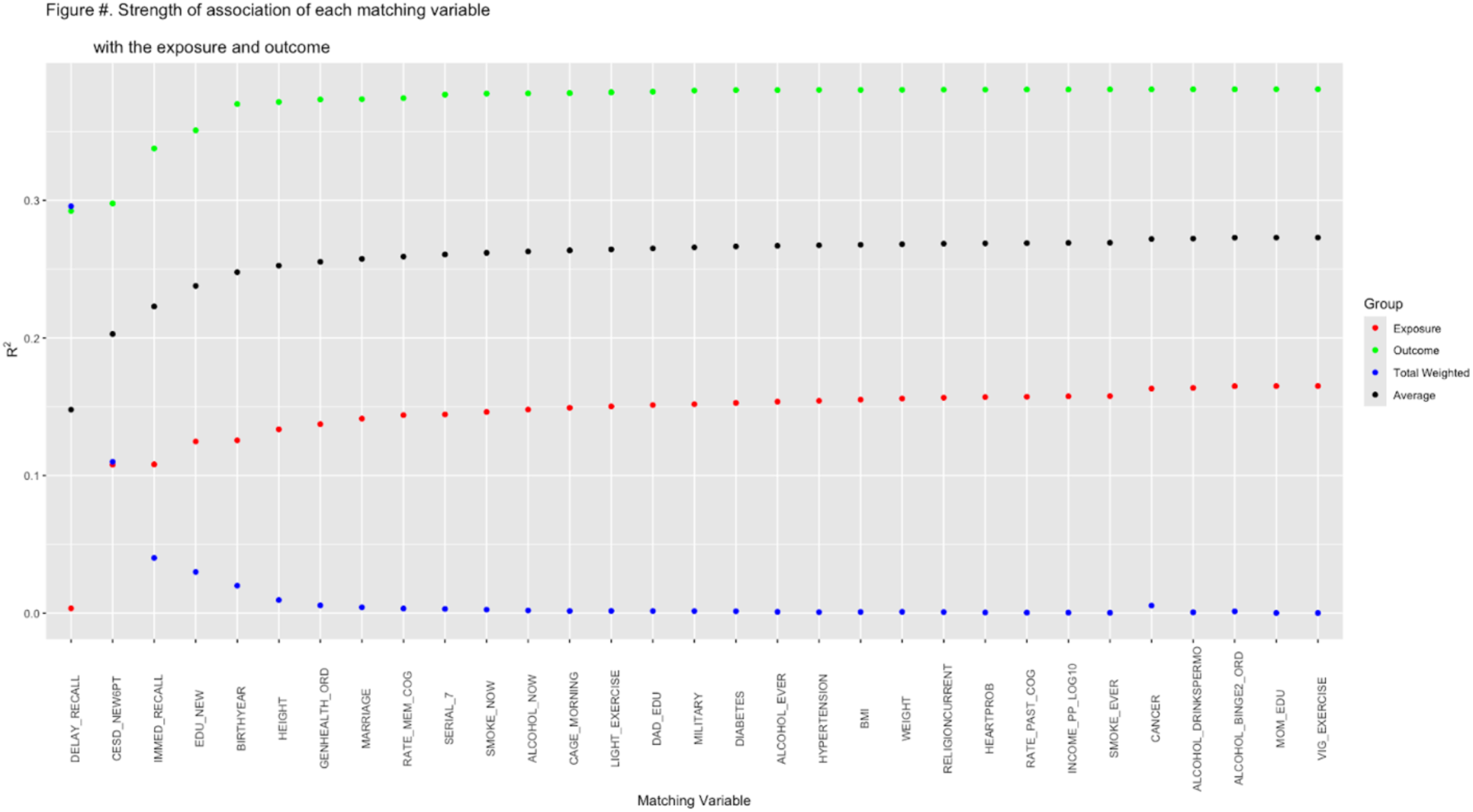
Incremental contribution of each variable included in exposure-covariate and outcome-covariate models to the sum of R^2 values from the two models.

**Appendix Figure 2:**
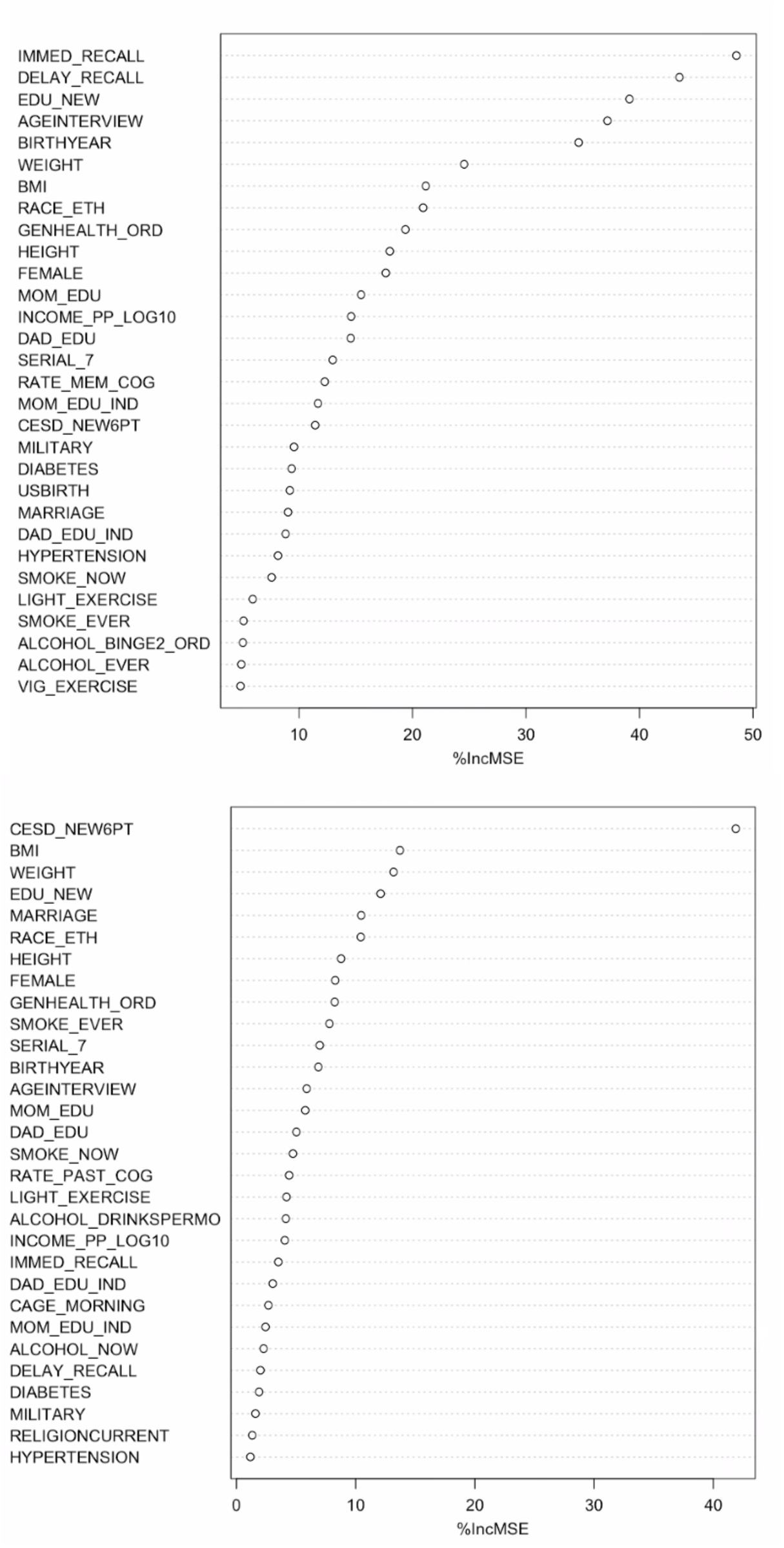
Mid-life variable importance (Mean Decrease in Accuracy) from random forest models predicting early life CESD in NLSY76 and later life cognition in HRS.

